# DermoExpert: Skin lesion classification using a hybrid convolutional neural network through segmentation, transfer learning, and augmentation

**DOI:** 10.1101/2021.02.02.21251038

**Authors:** Md. Kamrul Hasan, Md. Toufick E Elahi, Md. Ashraful Alam, Md. Tasnim Jawad

## Abstract

**Background and Objective:** Although automated Skin Lesion Classification (SLC) is a crucial integral step in computeraided diagnosis, it remains challenging due to inconsistency in textures, colors, indistinguishable boundaries, and shapes.

**Methods:** This article proposes an automated dermoscopic SLC framework named Dermoscopic Expert (DermoExpert). The DermoExpert consists of preprocessing and hybrid Convolutional Neural Network (hybrid-CNN), leveraging a transfer learning strategy. The proposed hybrid-CNN classifier has three different feature extractor modules taking the same input images, which are fused to achieve better-depth feature maps of the corresponding lesion. Those unique and fused feature maps are classified using different fully connected layers, which are then ensembled to predict the lesion class. We apply lesion segmentation, augmentation, and class rebalancing in the proposed preprocessing. We have also employed geometry- and intensity-based augmentations and class rebalancing by penalizing the majority class’s loss and combining additional images to the minority classes to enhance lesion recognition outcomes. Moreover, we leverage the knowledge from a pre-trained model to build a generic classifier, although small datasets are being used. In the end, we design and implement a web application by deploying the weights of our DermoExpert for automatic lesion recognition.

**Results:** We evaluate our DermoExpert on the ISIC-2016, ISIC-2017, and ISIC-2018 datasets, where the DermoExpert has achieved the area under the receiver operating characteristic curve (AUC) of 0.96, 0.95, and 0.97, respectively. The experimental results defeat the recent state-of-the-art by the margins of 10.0 % and 2.0 % respectively for the ISIC-2016 and ISIC-2017 datasets in terms of AUC. The DermoExpert also outperforms by a border of 3.0 % for the ISIC-2018 dataset concerning a balanced accuracy.

**Conclusion:** Since our framework can provide better-classification outcomes on three different test datasets, it can lead to better-recognition of melanoma to assist dermatologists. Our source code and segmented masks for the ISIC-2018 dataset will be publicly available for further improvements.

## 1. Introduction

### 1.1. Problem Presentation

Skin cancer, one in every three cancers worldwide [16], is a common type of cancer originating in the epidermis layer of the skin. Ultraviolet radiation exposure is one of the leading causes of skin cancer, roughly 90.0 % [52]. It was one of the five diseases in the United States (US) in a region under strong sunshine [58] in 2018. In 2019, there were 2490 females and 4740 males dropping their lives due to melanoma, whereas almost 20 people died in a day from melanoma in the US alone [81]. Age-standardized melanoma rates of the top twenty countries (see in Fig. 1) anticipated the disease rate, showing that a population would have melanoma if it had a standard age structure. The statistical results in Fig. 1 confirmed that the probability of having melanoma is more in males patients than in females subjects. Approximately 6850 new death cases due to melanoma were recorded in 2020, containing 4610 males and 2240 females [63]. However, a precise and robust early recognition is essential as the survival rate was as high as apparently 90.0 % in advance recognition [16].

**Figure 1:**
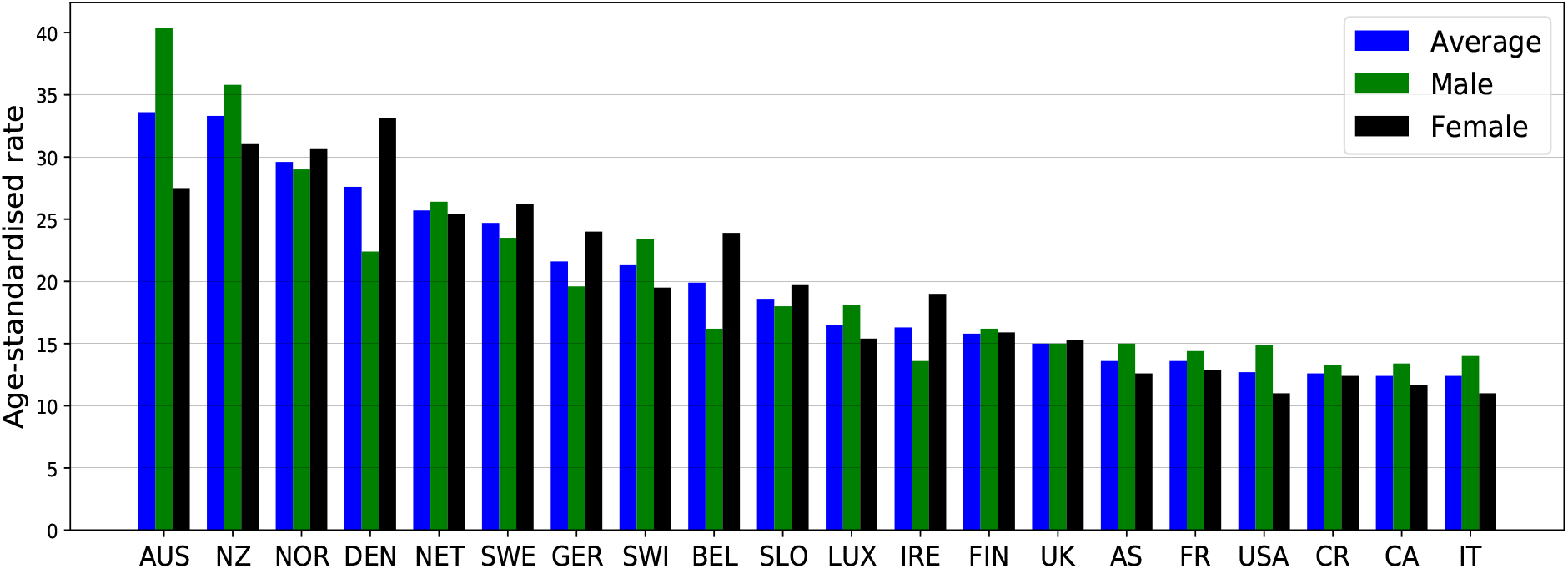
Age-standardized rates per 1.0 million of the top 20 countries with higher melanoma in 2018 [4]. The bars from left to right are for the decreasing melanoma case of the countries, such as Australia (AUS), New Zealand (NZ), Norway (NOR), Denmark (DEN), Netherlands (NET), Sweden (SWE), Germany (GER), Switzerland (SWI), Belgium (BEL), Slovenia (SLO), Luxembourg (LUX), Ireland (IRE), Finland (FIN), United Kingdom (UK) Austria (AS), France (FR), United States of America (USA), Czech Republic (CR), Canada (CA), Italy (IT)

Several imaging techniques, like confocal scanning laser microscopy, optical coherence tomography, ultrasound imaging, and dermoscopic imaging, are currently being practiced to diagnose skin cancer [64]. Dermoscopic images, also known as epiluminescence light microscopy, are most popularly utilized to investigate pigmented skin lesions by dermatologists [14]. Such a visual assessment, via the naked eye, may include a faulty-recognition, as it endures from the comparability between the lesions and healthy tissues [2, 33]. The dermatologists’ manual inspection is often tedious, time-consuming, and subjective; leading to different recognition results [2]. However, to mitigate all of the limitations mentioned earlier and improve skin cancer recognition preciseness, image-based Computer-aided Diagnosis (CAD) has been developed [32]. The classification action in an SLC-CAD system is a crucial component, which is a challenging task due to the presence of diverse artifacts, such as markers, body hair & fibers, air bubbles, reflections, on-uniform lighting, rolling lines, shadows, non-uniform vignetting, and patient-specific effects like lesion textures & colors, size of affected lesion area [24, 50]. Fig. 2 demonstrates some of those artifacts in the dermoscopic images.

**Figure 2:**
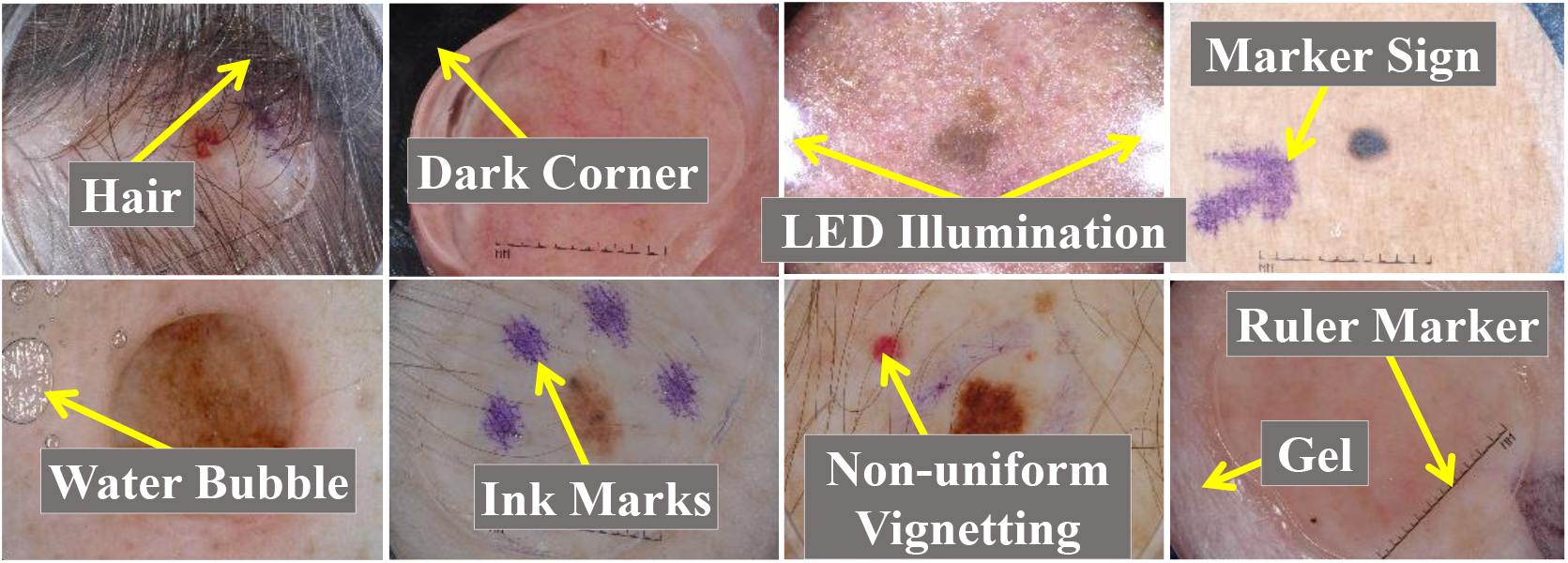
An example of the challenging dermoscopic images in ISIC dataset [10, 11, 20, 72] with different artifacts [24].

### 1.2. Related Works

Nowadays, several methods are being utilized for the SLC [8, 43]. We review and summarize numerous recent techniques, which are shortly described as follows: Yu et al. [77] proposed a novel CNN architecture for the SLC, where CNN learned from the multiple-image resolutions, leveraging pre-trained CNNs. They constructed a Fully Convolutional Residual Network (FCRN) and enhanced its capability by incorporating a multiscale contextual information scheme. The authors also integrated their proposed FCRN (for segmentation) and a deep residual network (for classification) to build a two-stage framework. Majtner et al. [47] developed an automatic melanoma detection system by employing the deep learning method, combining handcrafted RSurf features [48] and local binary patterns [1]. Finally, they applied Support Vector Machine (SVM) [15] for the SLC. A deep learning framework consisting of two FCRNs was proposed by Li and Shen [41] to get segmentation and coarse classification results concurrently. The authors also developed a Lesion Index Calculation Unit (LICU) to estimate the distance heat-map, where the coarse classification result was refined according to that generated distance map. Mahbod et al. [45] proposed an ensemble scheme, combining intra- and inter-architecture network fusion, where they used fine-tuning of AlexNet [37], VGGNet, and two variations of ResNets. The final prediction was achieved by adopting the SVM. Zhang et al. [80] aimed an Attention Residual Learning CNN (ARL-CNN) model for the SLC, coupling multiple ARL blocks, a global average pooling layer, and a classification layer. Each ARL block jointly handled residual and novel attention learning mechanisms to improve its ability for discriminating representation. Amin et al. [5] implemented the segmentation using the 2D wavelet transform and Ostu algorithm. They extracted lesion features using AlexNet and VGG16 models and employed a Principle Component Analysis (PCA) technique for the feature selection. Finally, the authors categorized the lesions using the k-nearest Neighbour (k-NN) [12, 23] and SVM. The effect of dermoscopic image size was analyzed by Mahbod et al. [46], using pre-trained CNNs and transfer learning. Three well-established CNNs, such as EfficientNetB0, EfficientNetB1, and SeReNeXt-50, were explored for the SLC. The authors also proposed and evaluated a multi-scale multi-CNN (MSM-CNN) fusion approach based on a three-level ensemble strategy that utilizes the three network architectures trained on cropped dermoscopic images of different scales. An architecture exploration framework was presented by Kwasigroch et al. [39] to identify the malignant melanoma. The hill-climbing search strategy was employed along with network morphism operations to explore the search space for finding a suitable network structure. Valle et al. [73] optimized the hyperparameter of the deep CNN models like ResNet-101-V2 [26] and Inception-V4 employing transfer learning and segmentation. They also performed extensive experiments to pick the best performing classifier using the ANOVA test. Finally, the authors concluded that the transfer learning and ensembling model is a better choice for designing the SLC systems. A Multi-class Multi-level (MCML) algorithm based on “divide and conquer” method was proposed by Hameed et al. [21]. The MCML consists of four integral parts: preprocessing, segmentation, feature extraction, and classification. However, their pipeline is highly dependent on preprocessing like hair, black frame, and circle removal. Gessert et al. [18] ensembled several deep learning methods, including EfficientNets, SENet [28], and ResNeXt [44], utilizing a selection strategy. The authors also fed the metadata as a feature vector, which was then concatenated with the CNN features. Khan et al. [35] developed a framework for the SLC, consisting of the localization of lesion ROI via faster region-based CNN, feature extraction, and feature selection by iteration-controlled Newton-Raphson method. Firstly, an artificial bee colony method was used for the contrast stretching, which was then used for the lesion segmentation. The DenseNet-201 was then utilized to extract the in-depth features. Finally, the authors employed a multilayer perceptron as a classifier. Almaraz-Damian et al. [3] proposed a pipeline, integrating preprocessing, feature extraction, feature fusion, and classification. As a preprocessing, the authors extracted ROIs of the lesion, where they also enhanced the intensity of the extracted ROIs. They extracted different handcraft features like shape, color, texture, and CNN features, where mutual information was employed to extract CNN features. Several classification methods, such as Linear Regression (LR), SVM, and Relevant Vector Machines (RVMs), were used to classify the ROIs. As a preprocessing, Mporas et al. [51] used a median filter, followed by bottom-hat filtering to detect the hair or similar noise. The segmentation was performed to extract lesion ROIs on the grayscale image using the active contour model. Finally, different color-based features were extracted, which were then classified using multilayer perceptron and different machine learning algorithms. Yilmaz and Trocan [76] compared the performance of deep CNN, such as AlexNet, GoogLeNet, and ResNet-50 for the SLC. The authors experimentally demonstrated that ResNet-50 was the best performing classifier, whereas AlexNet was better for time complexity. Pereira et al. [55] adopted the gradient-based histogram thresholding and local binary pattern clustering for skin lesions’ border-line characteristics. Then, border-line characteristics are concatenated with CNN to boost the lesion classification performance.

### 1.3. Our Contributions

While many methods are already developed and executed for the SLC, there is still room for performance enhancement. This article aims to build a robust and generic framework for the dermoscopic SLC, called DermoExpert. The preprocessing, the proposed hybrid-CNN classifier, and the transfer learning are the crucial integral steps of the intended SLC system. The proposed hybrid-CNN classifier employs a two-level ensembling. We have channel-wise concatenated 2D feature maps of different feature map generators to enhance the depth information in the first-level ensembling. Finally, we have aggregated the different outputs from fully-connected layers at the second-level ensembling. In the proposed preprocessing, we employ lesion segmentation, augmentation, and class rebalancing. The ROIs segmentation enables the classifier to learn only the specific lesion areas’ features while avoiding the surrounding healthy tissues. The precise segmentation, with less-coarseness, is a critical prerequisite step for the classification as it extracts abstract region and detailed structural description of various types of skin lesions. The segmentation has performed by using our recent state-of-the-art DSNet [24] after fine-tuning with other ISIC datasets. We utilize geometry- and intensity-based image augmentation to enhance training image samples and evade overfitting. We also use a class rebalancing by combining additional images from the ISIC archive and weighting the loss function to tackle the unwanted biasing towards the majority class. Moreover, the transfer learning was applied to all the different feature extractors in the proposed hybrid classifier. To our best knowledge, our proposed DermoExpert has achieved state-of-the-art results on the three IEEE International Symposium on Biomedical Imaging (ISBI) datasets, such as ISIC-2016, ISIC-2017, and ISIC-2018 with respectively two, three, and seven lesion classes, while being an end-to-end system for the SLC. Additionally, we have implemented and compared the performance of our DermoExpert against several well-established deep learning classification approaches like Xception, ResNet, and DenseNet under the same experimental environments and preprocessing using the same dataset. Finally, we have implemented a web application by deploying the trained DermoExpert for the clinical application, which runs in a web browser.

The remaining sections of the article are organized as follows: section 2 explains the proposed methodologies and datasets, where we explicitly mention the extensive experiments. Section 3 describes the obtained results from different extensive experiments. The results are explained with a proper interpretation in section 4, where we also present a user application by employing the trained model for the SLC. Finally, section 5 concludes the article with future works.

## 2. Methods and Materials

This section focuses on methods and materials utilized for the study, where subsection 2.1 explains the proposed DermoExpert and datasets for the SLC. In subsection 2.2, we present the metrics for evaluation and hardware used to conduct this research. Subsection 2.3 presents the training protocol and experimental details.

### 2.1. Proposed Framework

The proposed DermoExpert consists of dermoscopic image preprocessing, transfer learning, and a proposed hybrid-CNN classifier (see in Fig. 3). We have employed lesion segmentation for ROI extraction, augmentation, and class rebalancing as a preprocessing. We validate our DermoExpert with three different datasets having a different number of target classes to provide the evidence of generality and versatility of the proposed DermoExpert. There are three distinct inputs in the DermoExpert (see in Fig. 3) for three different datasets, which are termed as *I*_1_, *I*_2_, and *I*_3_. The input *I*_1_, *I*_2_, or *I*_3_ generates the corresponding output *O*_1_, *O*_2_, or *O*_3_ using the proposed preprocessing and hybrid-CNN classifier. The different integral and crucial parts of the DermoExpert are briefly described as follows:

**Figure 3:**
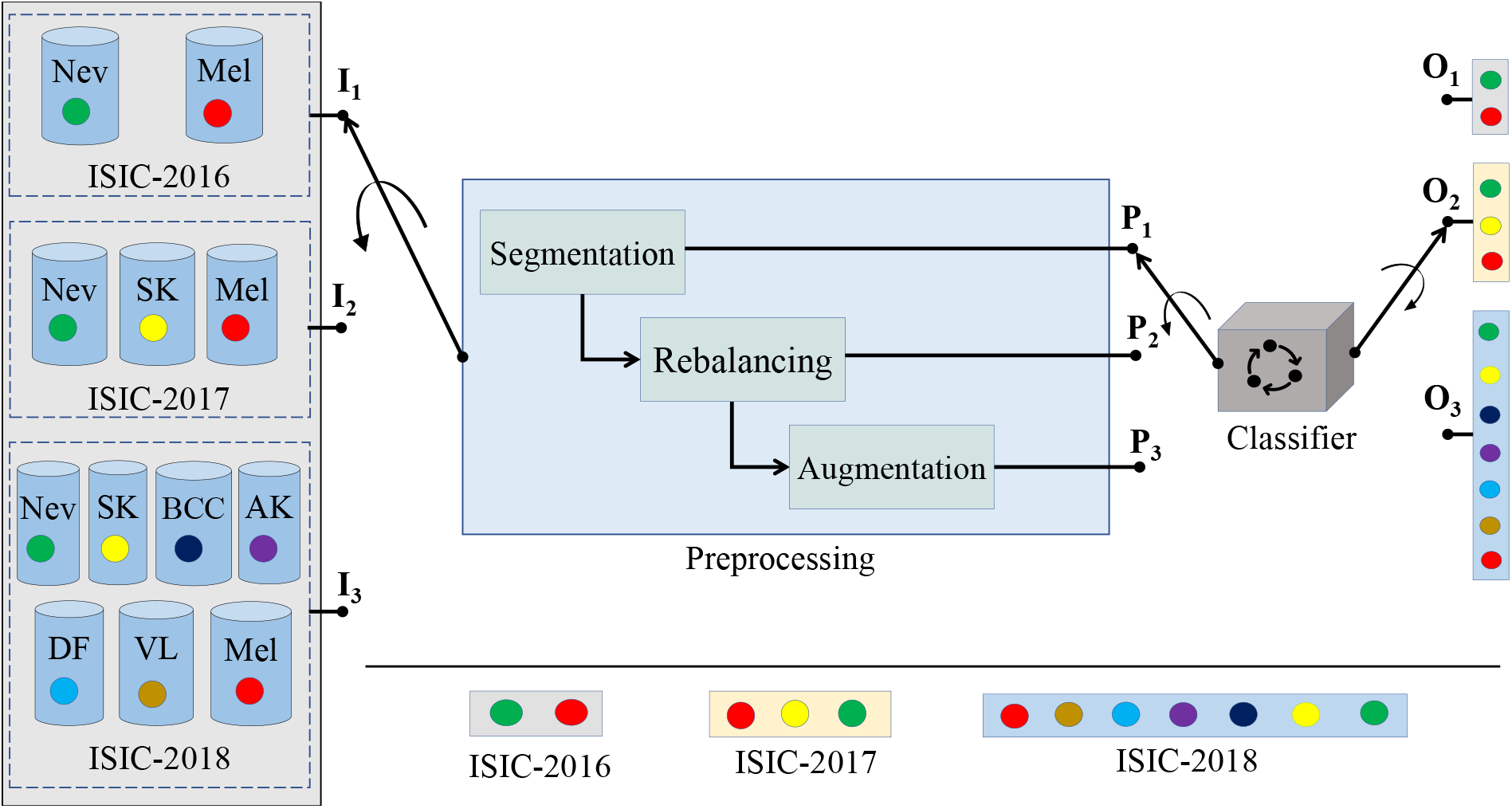
The proposed DermoExpert for the SLC, where the preprocessing has been incorporated with the pipeline to build a precise and robust diagnostic system. The input *I*_1_, *I*_2_, or *I*_3_ is followed by the preprocessing (*P*_1_, *P*_2_, or *P*_3_) and then by the proposed classifier to generate the corresponding output *O*_1_, *O*_2_, or *O*_3_.

#### 2.1.1. Datasets

The three well-known datasets of skin lesions, such as ISIC-2016 [20], ISIC-2017 [11], and ISIC-2018 [10, 72] are utilized to evaluate the proposed DermoExpert (see Fig. 3) respectively denoting by *I*_1_, *I*_2_, and *I*_3_. Table 1 manifests the class-wise distributions and short descriptions of the utilized datasets. The ISIC-2016 is a binary classification task to distinguish the lesions as either Nevus (Nev) or Melanoma (Mel). The ISIC-2017 and ISIC-2018 are the multi-class categorization tasks. In the ISIC-2017, the lesion requires to classify as either Nevus (Nev) or Seborrheic keratosis (SK) or Melanoma (Mel). The ISIC-2018 comprises of Nevus (Nev), Seborrheic keratosis (SK), Basal Cell Carcinoma (BCC), Actinic Keratosis (AK), Dermatofibroma (DF), Vascular Lesion (VL), and Melanoma (Mel) classes. However, the ground-truths of validation and test images are not provided for the ISIC-2018 dataset. We have applied a cross-validation technique for ISIC-2018 dataset to choose training, validation, and testing images. The resolutions in pixels of all 8-bit dermoscopic images are 540 *×* 576 to 2848 *×* 4288, 540 *×* 576 to 4499 *×* 6748, and 450 *×* 600 respectively for ISIC-2016, ISIC-2017, and ISIC-2018 datasets. The class-distribution of all ISIC datasets (see in Table 1) confirms that the images are imbalanced, which makes the classifier to be biased towards the particular class holding more samples. However, we have employed several rebalancing techniques to build a generic classifier for the skin lesion diagnosis even though datasets are imbalanced.

**Table 1:** A concise description and class-distribution of the ISIC-2016, 2017, and 2018 datasets.

| SL # | Class Types | Description | Train | Validation | Test |
| --- | --- | --- | --- | --- | --- |
| 01 | Nevus (Nev) | Benign tumor derived from melanocytic | 727 | * | 304 |
| 02 | Melanoma (Mel) | Malignant tumor derived from melanocytic | 173 | * | 75 |
| Total Images in ISIC-2016 |  |  | 900 | * | 379 |
| 01 | Nevus (Nev) | Benign tumor derived from melanocytic | 1372 | 78 | 393 |
| 02 | Seborrheic Keratosis (SK) | Benign tumor derived from non-melanocytic | 254 | 42 | 90 |
| 03 | Melanoma (Mel) | Malignant tumor derived from melanocytic | 374 | 30 | 117 |
| Total Images in ISIC-2017 |  |  | 2000 | 150 | 600 |
| 01 | Nevus (Nev) | Benign tumor derived from melanocytic | 6705 | * | * |
| 02 | Seborrheic Keratosis (SK) | Benign tumor derived from non-melanocytic | 1099 | * | * |
| 03 | Basal Cell Carcinoma (BCC) | Benign tumor derived from nonmelanocytic | 514 | * | * |
| 04 | Actinic Keratosis (AK) | Benign tumor derived from keratinocytes | 327 | * | * |
| 05 | Dermatofibroma (DF) | Benign tumor derived from histiocytes | 115 | * | * |
| 06 | Vascular Lesion (VL) | Benign tumor derived from blood vessel cell | 142 | * | * |
| 07 | Melanoma (Mel) | Malignant tumor derived from melanocytic | 1113 | * | * |
| Total Images in ISIC-2018 |  |  | 10015 | * | * |

\*: Not available publicly

#### 2.1.2. Proposed Preprocessing

The proposed preprocessing in Fig. 3 consists of segmentation, augmentation, and class rebalancing, which are briefly described as follows:

##### Segmentation

The segmentation, to separate homogeneous lesion areas, is the critical component for diagnosis and treatment pipeline [27]. It is also a crucial prerequisite for the skin lesion diagnosis as it extracts promising skin lesion features and delivers critical information about the shapes and structures. A recent state-of-the-art DSNet [24] for dermoscopic skin lesion (ISIC-2017) segmentation has been adopted as a lesion ROI extractor. We fine-tune the selected DSNet with the ISIC-2016 and ISIC-2018 datasets to extract the lesion ROI of all three datasets as it was trained and tested on only the ISIC-2017 dataset.

##### Augmentation

CNNs are heavily reliant on big data to avoid overfitting. Unfortunately, many application domains, such as automation in lesion disease diagnosis, suffer from a small size as a massive number of manually annotated training images are not yet available [22]. Augmentation is a very potential preprocessing for training the deep learning models as they are highly discriminative [30]. Data augmentation encompasses a technique that enhances training datasets’ size and quality to build a better-CNN classifier [62]. The geometry-based augmentation, such as a rotation (around *row/*2 and *col/*2) of 180*°* and 270*°*, and the intensity-based augmentations, such as gamma, logarithmic, & sigmoid corrections, and stretching, or shrinking the intensity levels are employed in the proposed preprocessing. The gamma correction with two gammas (*γ*) of 0.7 and 1.7 has performed to change the luminance of the dermoscopic images by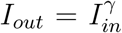, where *I*_*out*_ and *I*_*in*_ are the output and input luminance values. The logarithmic correction, for enhancing an image to provide better contrast and a more structural detailed image, has employed by *I*_*out*_ = *G ×* log (1 + *I*_*in*_), where *G* = 0.5 and *I*_*out*_ & *I*_*in*_ are the gain and input & output images, respectively. We have employed sigmoid correction by 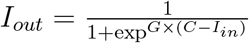, where G = 15,*C* = 0.4, and *I*_*out*_ & *I*_*in*_ are the multiplier in exponential’s power, cutoff of the function that shifts the characteristic curve in horizontal direction, and input & output images, respectively. We also stretch or shrink the intensity levels between the minimum and the maximum intensities.

### Rebalancing

All of the three dermoscopic datasets are imbalanced, as presented in Table 1. This scenario is quite common in the medical diagnosis field as manually annotated training images are not adequately available [22], where the positive cases are the minority compared to negative cases. The unwanted biasing towards the majority class is likely to happen in the supervised CNN-based classifiers. However, we have joined additional images from the ISIC archive website [31], and we also penalize the majority class by weighting the loss function. Such a weighting pays more attention to the underrepresented class. Here, we improve the weight of samples from underrepresented classes with a factor of *W*_*j*_ = *N*_*j*_*/N*, where *W*_*j*_, *N*, and *N*_*j*_ are the weight for class *j*, the total number of samples, and the number of samples in class *j*, respectively.

### Preprocessing Employment

We have sub-divided the preprocessing (*P*) into the segmentation (*P*_1_), segmentation & class rebalancing (*P*_2_), and segmentation, class rebalancing, & augmentation (*P*_3_) to investigate their effects in the proposed DermoExpert for the SLC.

#### 2.1.3 Proposed CNN-based Classifier and Transfer Learning

Deep CNNs are an excellent feature extractor, avoiding complicated and expensive feature engineering. It has achieved tremendous success since 2012 [57]. Sometimes, it rivals human expertise. For example, CheXNet [56] was trained on more than 1.0 million chest X-rays. It was capable of achieving a better performance than the four experts. Again, Kermany et al. [34] trained Inception-V3 network with roughly 1.0 million optical coherence tomographic images. They compared the results with six radiologists, where experts got high sensitivity but low specificity, while the CNN-based system acquired high values of balanced accuracy. However, those above methods are blessed with a massive number of labeled images, which are hard to collect as it needs much professional expertise for annotation [75]. Moreover, individual CNN architecture may have different capabilities to characterize or represent the image data, which is often linked to a network’s depth [38]. However, CNN’s maybe indirectly limited when employed with highly variable and distinctive image datasets with limited samples, such as dermoscopic image datasets [10, 11, 20, 72]. In this context, we propose a hybrid-CNN classifier by leveraging several core techniques of the current CNN networks to build a generic skin lesion diagnostic system with limited training images. Fig. 4 depicts the proposed hybrid-CNN classifier. In our hybrid-CNN, the input batch of images is simultaneously passed through three different Feature Map Generators (FMGs) to obtain different presentations of the feature maps (see the output of the encoders in Fig. 4). The proposed hybrid-CNN classifier comprises the following integrated parts:

**Figure 4:**
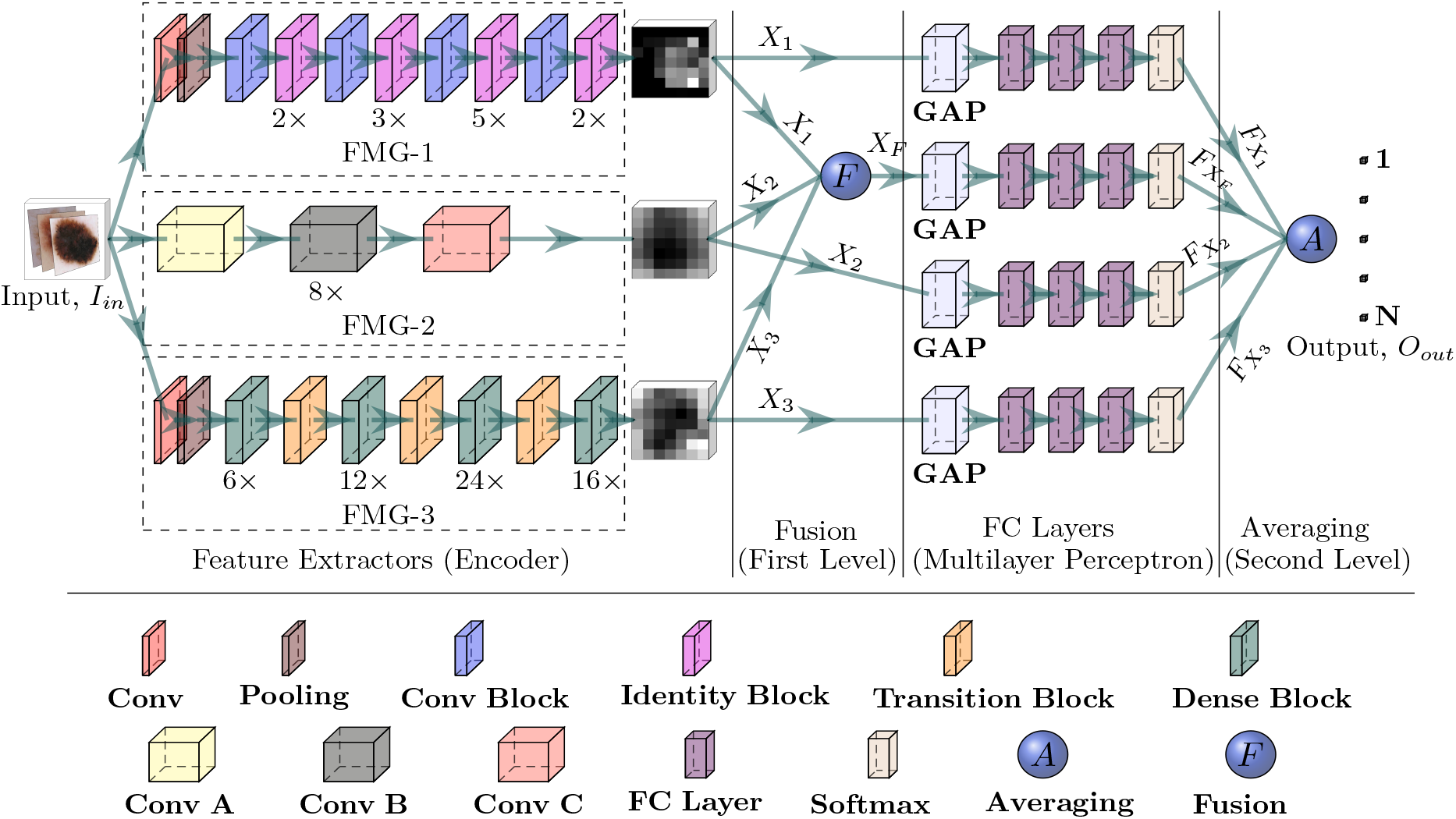
The proposed hybrid-CNN classifier, where three different feature extractors, also called an en-coder, receives the same input image (*I*_*in*_). The first step, encoders, are followed by the second step, called a fusion (first-level ensembling). Then, the third step, called the FC layer, is followed by the fourth step, called an averaging (second-level ensembling), to get the final output (*O*_*out*_).

#### Feature Map Generators-1 (FMG-1)

The FMG-1 (*f* ^1^) consists of the identity or residual and convolutional blocks [25], where the skip connections allow the information to flow or skip. Fig. 5 shows the constructional details of the identity and convolutional blocks. The skip connection in the residual blocks, as in the proposed hybrid-CNN classifier, has two benefits: firstly, the new layers will not hamper the performance as regularisation will skip over them, and secondly, if the new layers are useful, even with the presence of regularisation, the weights or kernels of the layers will be non-zero. However, a 7 *×* 7 input convolution is applied in FMG-1 before the identity and convolutional blocks, followed by a max-pooling with a stride of 2 and a pool size of 3 *×* 3. By stacking these blocks on top of each other (see Fig. 4), an FMG-1 has been formed to get the feature map, where the notation (*n×*) under the identity block denotes the number of repetitions (*n* times). The output feature map from the FMG-1 is defined as *X*_1_ = *f* ^1^(*I*_*in*_), where *X*_1_ *∈ ℛ*^*B×H×W ×D*^, and *B, H, W*, and *D* respectively denote the batch size, height, width, and depth (channel).

**Figure 5:**
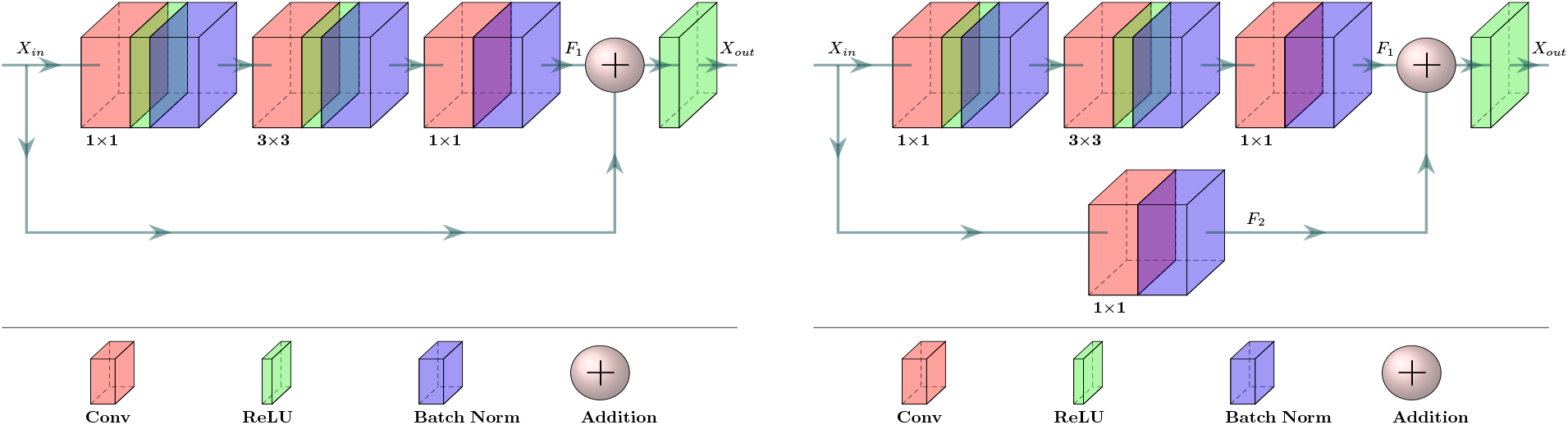
The residual (or identity) (left) and convolutional (right) blocks [25] of the FMG-1. The output (*X*_*out*_) is the summation of *X*_*in*_ and the process (*F*), where *X*_*out*_ = *F*_1_(*X*_*in*_) + *X*_*in*_ for residual block and *X*_*out*_ = *F*_1_(*X*_*in*_) + *F*_2_(*X*_*in*_) for convolutional block.

#### Feature Map Generators-2(FMG-2)

The FMG-2 (*f* ^2^) consists of the entry flow (Conv A), middle flow (Conv B), and exit flow (Conv C) blocks, which were originally proposed by Chollet [9]. The constructional details of those blocks are given in Fig. 6. The batch of images first goes through the entry flow, then through the middle flow, which is repeated eight times (8*×*), and finally through the exit flow. As in the proposed hybrid-CNN classifier, all the flows have used depth-wise separable convolution and residual connection. The former one is used to build a lightweight network, whereas the latter one for the benefits, which were mentioned earlier. The output feature map from the FMG-2 is defined as *X*_2_ = *f* ^2^(*I*_*in*_), where *X*_2_ *∈ ℛ*^*B×;H×W×D*^, and *B, H, W*, and *D* respectively denote the batch size, height, width, and depth (channel).

**Figure 6:**
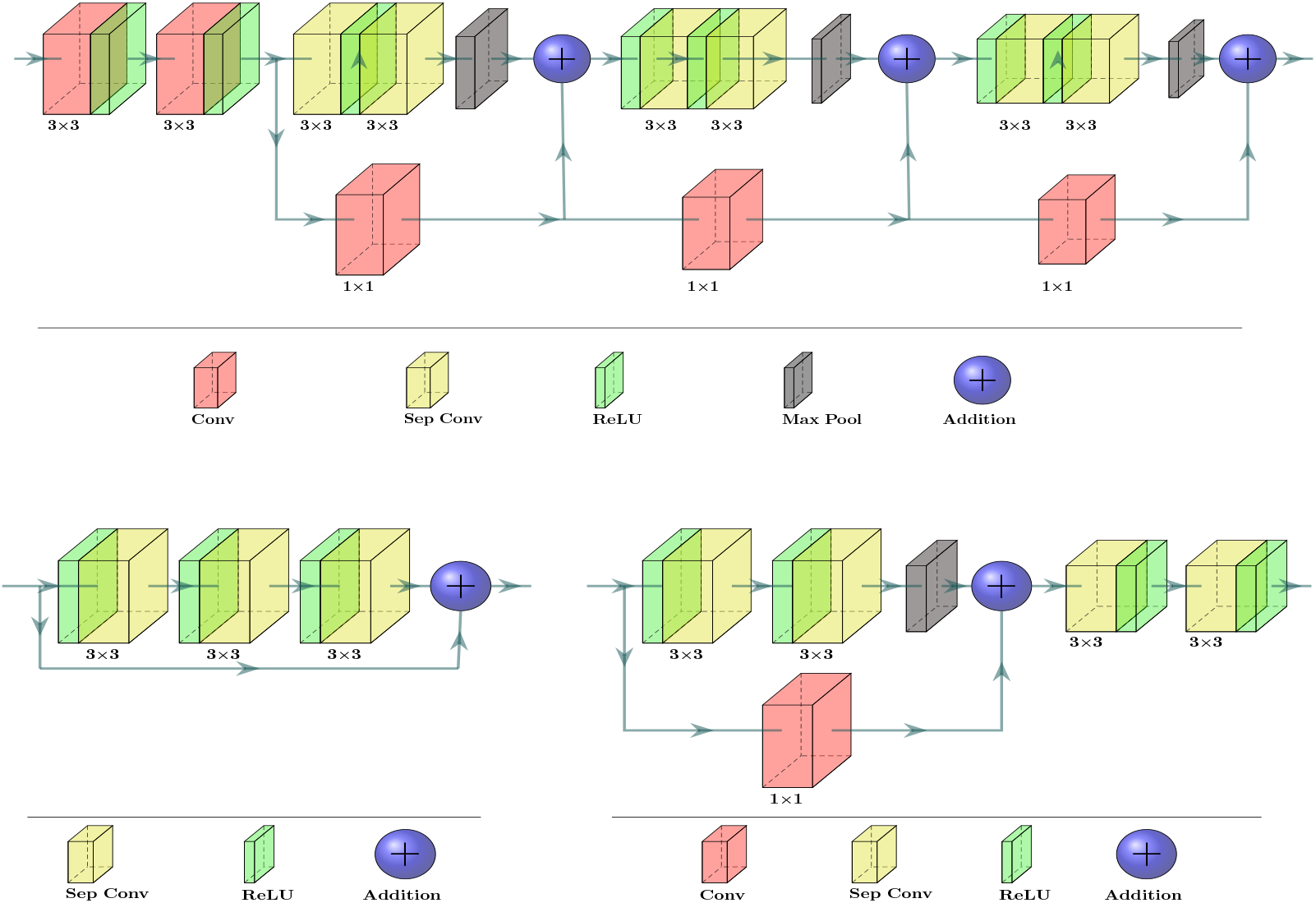
The entry flow (Conv A) (top), middle flow (Conv B) (bottom-left), and exit flow (Conv C) (bottom-right) blocks [9] of the FMG-2, where depth-wise separable convolutions were employed in lieu of traditional convolutions to make it lightweight for real-time applications.

#### Feature Map Generators-3 (FMG-3)

The remaining FMG-3 (*f* ^3^) consists of the dense and transition blocks [29], where the constructions of those blocks are displayed in Fig. 7. The FMG-3, as in the proposed hybrid-CNN classifier, gets rid of the requirement of learning repetitive features, which can learn the absolute features of the skin lesion [24]. The feature re-usability of FMG-3 reduces the vanishing-gradient problem and strengthens the feature propagation [29]. It also enables the convolutional layer to access the gradients of all the previous layers by using a skip connection, as depicted in Fig. 7. However, the output feature map from the FMG-3 is defined as *X*_3_ = *f* ^3^(*I*_*in*_), where *X*_3_ *∈* ℛ^*B×H×W×D*^, and *B, H, W*, and *D* respectively denote the batch size, height, width, and depth (channel).

**Figure 7:**
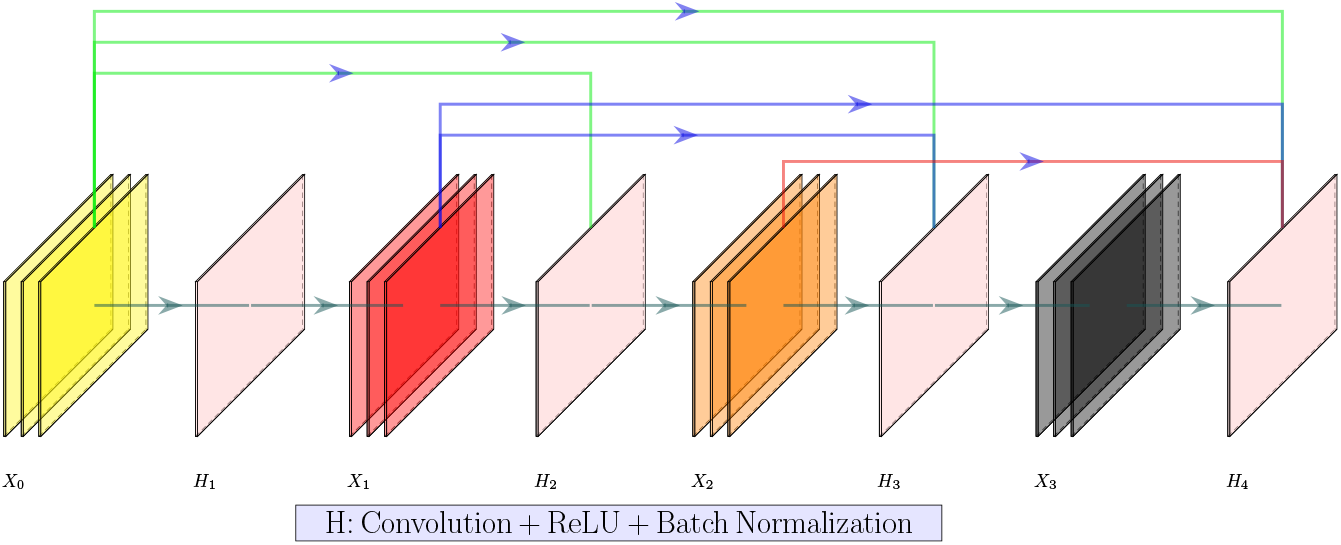
Typical dense block of the FMG-3 for a growth rate of 3.0. Each *n*^*th*^ layer of a dense block accepts all previous convolutional layers’ feature maps. The mathematical expression [29] of such a reusibiility is *X*_*n*_ = *H*_*n*_([*X*_0_, *X*_1_, *X*_2_, …, *X*_*n−*1_]), where *H*_*n*_ is the composite function [29] of the *n*^*th*^ layer, which consists of a convolution, ReLU, and batch normalization.

#### Fusion (First-level ensembling)

We have employed those three FMGs, to get the distinct feature maps (see in Fig. 8 (c), Fig. 8 (d), and Fig. 8 (e)) for building a proposed hybrid-CNN classifier. It is very impractical to choose a better-fusion mechanism without experiments as there are many ways to get a fused map (*X*_*F*_). However, we perform two types of fusion, such as fusion by channel-concatenation (see Fig. 8 (g)) and fusion by channel-averaging (see Fig. 8 (f)), and we have named them as a first-level ensembling. In channel-concatenation, the fused feature map is 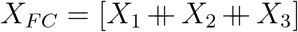, where *X*_*F C*_ *∈*; ℛ ^*B×H×W ×*3*D*^ and **Σ** denotes the channel concatenation. In channel-averaging, the fused feature map is 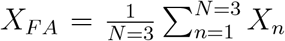, where *X*_*F A*_ *∈* ℛ ^*B×H×W ×D*^, Σ is the element-wise summation, and *N* is the numbers of FMG.

**Figure 8:**
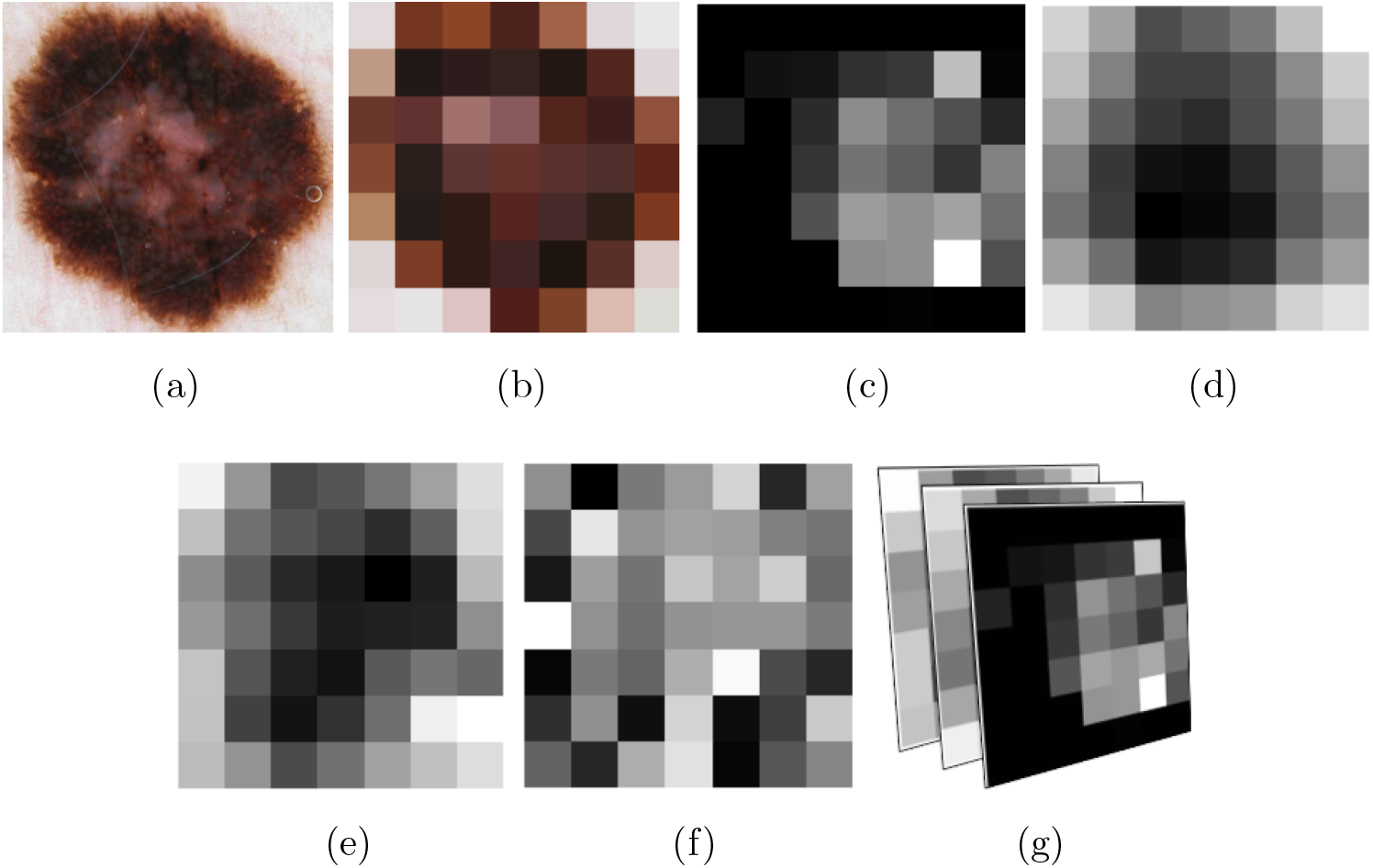
The extracted feature map from different FMGs with corresponding original input image (a), where (b) the down-sampled (7 *×* 7) image using nearest-neighbor interpolation, (c) the output of FMG-1, (d) the output of FMG-2, (e) the output of FMG-3, (f) the average of the three FMGs, and (g) the channel-wise concatenation of the output of the three FMGs.

#### Fully Connected Layer

The different feature maps are classified into desired categories using the Fully Connected (FC) layers, where the output is denoted by *F*_*M*_ for *M*^*th*^ input feature map. However, to vectorize the 2D feature maps into a single long continuous linear vector before the FC layer, we use a Global Average Pooling (GAP) layer [42], which improves generalization and prevents overfitting. The GAP layer performs a more extreme dimensionality reduction to avoid overfitting. An *height × weight × depth* dimensional tensor, in GAP, is reduced to a 1 *×* 1 *× depth* vector by transferring *height × width* feature map to a single number. Such a GAP layer also contributes to the lightweight design of the CNN classifiers. Additionally, each FC layer is followed by a dropout layer [67] as a regulariser, where we randomly set 50.0 % neurons of the FC layer to zero during the training. Such a dropout layer assists in building a generic CNN classifier by reducing the overfitting.

#### Averaging (Second-level ensembling)

Finally, the output probability (*O*_*j*=1,2,3_) is the average of the outputs of different *F*_*M*_, and we have named it as a second-level ensembling. The output (*O*_*j*=1,2,3_) lies in *N* -dimensional space, where *O*_1_ *∈ ℛ*^*N* =2^, *O*_2_ *∈ ℛ*^*N* =3^, and *O*_3_ *∈ ℛ*^*N* =7^ respectively for the inputs *I*_1_, *I*_2_, and *I*_3_ (see subsection 2.1 and Fig. 3) by applying the proposed preprocessing (either *P*_1_ or *P*_2_ or *P*_3_).

#### Possible hybrid-CNN classifiers

However, this article proposes five possible ensembling classifiers by using first- and second-level ensembling, which are enlisted as follows:

1. **Method-1:** Selection of only the first-level ensembling by using the fused feature map *X*_*F C*_, and performing the classification using an FC layer 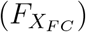for achieving a final probability *O*_*j*_
2. **Method-2:** Selection of only the first-level ensembling by using the fused feature map *X*_*F A*_, and performing the classification using an FC layer 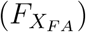 for achieving a final probability *O*_*j*_
3. **Method-3:** Selection of only the second-level ensembling by employing the feature maps (*X*_1_, *X*_2_, and *X*_3_) except the fused maps (*X*_*F C*_ and *X*_*F A*_), and performing the classification using the FC layers 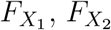, and 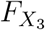) for achieving a final probability *O*_*j*_
4. **Method-4:** Selection of both the first and second-level ensembling by using the feature maps (*X*_1_, *X*_2_, *X*_3_, and *X*_*F A*_) except the fused map (*X*_*F C*_), and performing the classification using the FC layers 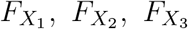, and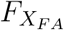) for achieving a final probability *O*_*j*_
5. **Method-5:** Selection of both the first- and second-level ensembling by using the feature maps (*X*_1_, *X*_2_, *X*_3_, and *X*_*F C*_) except the fused map (*X*_*F A*_), and performing the classification using the FC layers (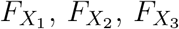 and 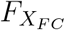) for achieving a final probability *O*_*j*_ However, we perform the ablation studies (see in subsection 3.3) on those five different methods mentioned above to achieve the best hybrid-CNN classifier for the SLC.

#### Transfer Learning

Moreover, when the data number is relatively small, as the skin lesion datasets in this article, the model overfit after several epochs. However, the scarcity of such relatively small medical image datasets has been partially overcome by employing a transfer learning [61, 68]. It applies the representations learned by a previous model and employs a new domain, reducing the need for sizeable computational power [69]. However, in the proposed DermoExpert, we use the previously trained weights to our FMG-1, FMG-2, and FMG-3 for transferring the knowledge.

### 2.2. Hardware and Evaluation

#### Hardware

We have implemented our DermoExpert on a *Windows-*10 machine using the Python programming language with different Python and Keras [17] APIs. The hardware configuration of the used machine are: Intel^*(r)*^ Core^TM^ i7-7700 HQ CPU @ 2.80 *GHz* processor with Install memory (RAM): 16.0 *GB* and GeForce GTX 1060 GPU with 6 *GB* GDDR5 memory.

#### Evaluation Metrics

We utilize recall, specificity, and intersection over union (IoU) for measuring the segmentation performance quantitatively. The recall and specificity measure the percentage of true and wrong regions, whereas the IoU measures the overlap between the true and predicted masks. We evaluate the proposed SLC results using the recall, precision, and F1-score. The recall quantifies the type-II error (the lesion, with the positive syndromes, inappropriately fails to be nullified), and precision quantifies the positive predictive values (percentage of truly positive recognition among all the positive recognition). The F1-score indicates the harmonic mean of recall and precision, conferring the tradeoff between them. We have also reported the confusion matrix for evaluating the DermoExpert by investigating the class-wise performance of the SLC. Moreover, the Receiver Operating Characteristics (ROC) with Area Under the ROC Curve (AUC) value is also used to quantify any randomly picked sample’s prediction probability.

### 2.3. Training Protocol and Experiments

As we segment the lesion using the recent state-of-the-art DSNet, all the kernels have been initialized with the pre-trained weights of the DSNet. We resize all the images to 192 *×* 256 pixels using the nearest-neighbor interpolation for the segmentation, as the DSNet receives the images having a resolution of 192 *×* 256 pixels. Additionally, we have standardized and rescaled the training images to [0 1]. The fine-tuning of the DSNet has been conducted using the following loss function (*L*) [24] as Eq. 1.

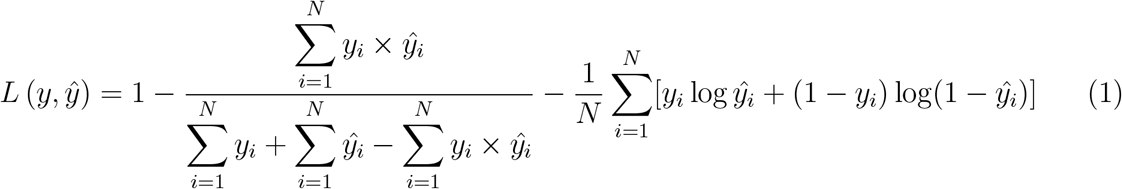

where *y* and *ŷ, N* respectively denote the true and predicted label, the total number of and pixels.

The pre-trained weights from ImageNet [13] were applied to initialize the kernels of all the FMGs. Xavier distribution, also called glorot uniform distribution [19], is used to initialize the kernels in FC layers. It draws the samples from a truncated normal distribution centered on 0.0 with a standard deviation of 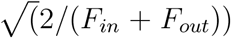. *F*_*in*_ and *F*_*out*_ respectively denotes the number of input and output units in the weight tensor. The aspect ratio distribution extracted ROIs of the lesion for ISIC-2016, ISIC-2017, and ISIC-2018 datasets reveal that most of the ROIs have an aspect ratio of 1 : 1. Therefore, all the ROIs have been resized to 192 *×* 192 pixels using a nearest-neighbor interpolation for classification using the Dermo Expert. The categorical cross-entropy function is used as a loss function in our Dermo Expert. However, it is very impractical to guesstimate the useful optimizer and the learning rate as they are highly dependent on the networks and datasets. In this literature, we perform extensive experiments for selecting those hyperparameters as described in subsection 3.2. We set the initial epochs as 200 and stop the training using a callback function when the validation loss has stopped improving. However, we conduct several extensive experiments to achieve the highest possible performance for a robust and accurate SLC system. Firstly, we select the optimizer with learning rate and the best hybrid-CNN classifier (see in sub-section 2.1.3), for the proposed DermoExpert, via comprehensive experiments. Then, we perform the following experiments:

1. We fine-tuned DSNet on the ISIC-*n* dataset, where *n* = 2016, 2017, 2018. Then, we extract the lesion ROI and resize the images to 192 *×* 192
2. We complete the classification using the proposed DermoExpert on those segmented ROIs
3. We rebalance the lesion classes of the segmented ROIs since the class distributions are imbalanced, and then do the classification
4. Finally, we add the intensity- and geometry-based augmentations on the experiment-3

We have repeated all the experiments mentioned above for the three different datasets, such as ISIC-2016, ISIC-2017, and ISIC-2018.

## 3. Experimental Results

This section reports the qualitative and quantitative results through several extensive experiments. The segmentation results of the fine-tuned DSNet and the qualitative results for different augmentations are detailed in subsection 3.1. We present ablation studies for optimizer and learning rate adaptation in subsection 3.2. We also exhibit the ablation examinations in subsection 3.3 for the best hybrid-CNN classifier selection (see in subsection 2.1.3) for the DermoExpert. The classification results on ISIC-2016, ISIC-2017, and ISIC-2018 datasets are manifested in the subsections 3.4, 3.5, and 3.6, respectively. Finally, we compare our results with the state-of-the-art results for the SLC in subsection 3.7.

### 3.1. Segmentation and Augmentation

Table 2 displays the quantitative results for the lesion ROI extraction (segmentation) for further classification in the SLC. The results in Table 2 demonstrate that the type-I errors are 3.8 %, 6.5 %, and 4.6 % respectively for ISIC-2016, ISIC-2017, and ISIC-2018 datasets, whereas the respective type-II errors are 9.2 %, 12.0 %, and 8.9 %. Such fewer type-I and type-II errors reveal that our segmented ROIs are blessed with fewer false-negative and false-positive regions, making them better-applicable for the SLC. The qualitative results of the segmented masks (see in Fig. 9) depict that the extracted green ROIs have approximately coincided with the actual yellow ROIs. More segmentation results for all the test images are available on YouTube (ISIC-2016^2^, ISIC-2017^3^, and ISIC-2018^4^). The qualitative results in Fig. 9 also confirm that the segmented masks are also as better as in the microscopic skin images (first and last columns of the second row). The quantitative and qualitative representations of the segmentation results demonstrate that the mean overlapping between the true and predicted masks of all the test images is as high as the recent state-of-the-art results for dermoscopic lesion segmentation [6, 71, 74, 82]. However, all the segmented ROIs are further processed for the augmentation as both the quantitative and qualitative results of the segmentation point that they yield the most reliable ROIs for the SLC.

**Table 2:**
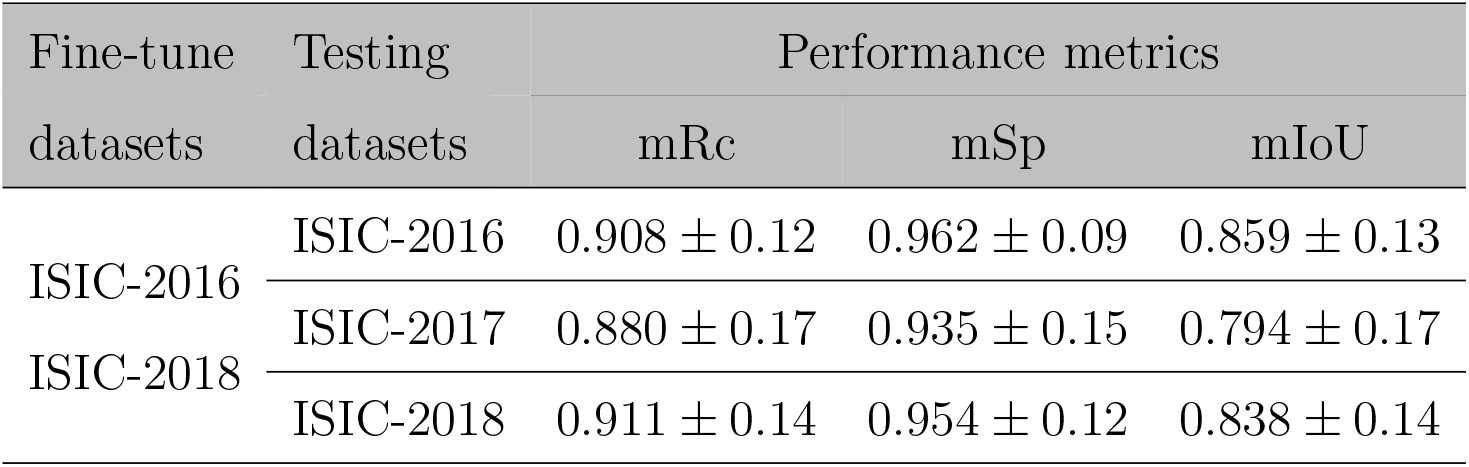
Segmentation results on the test datasets of the ISIC-2016, ISIC-2017, and ISIC-2018 from the fine-tuned DSNet, where the mRc, mSp, and mIoU respectively indicate the mean recall, specificity, and IoU.

**Figure 9:**
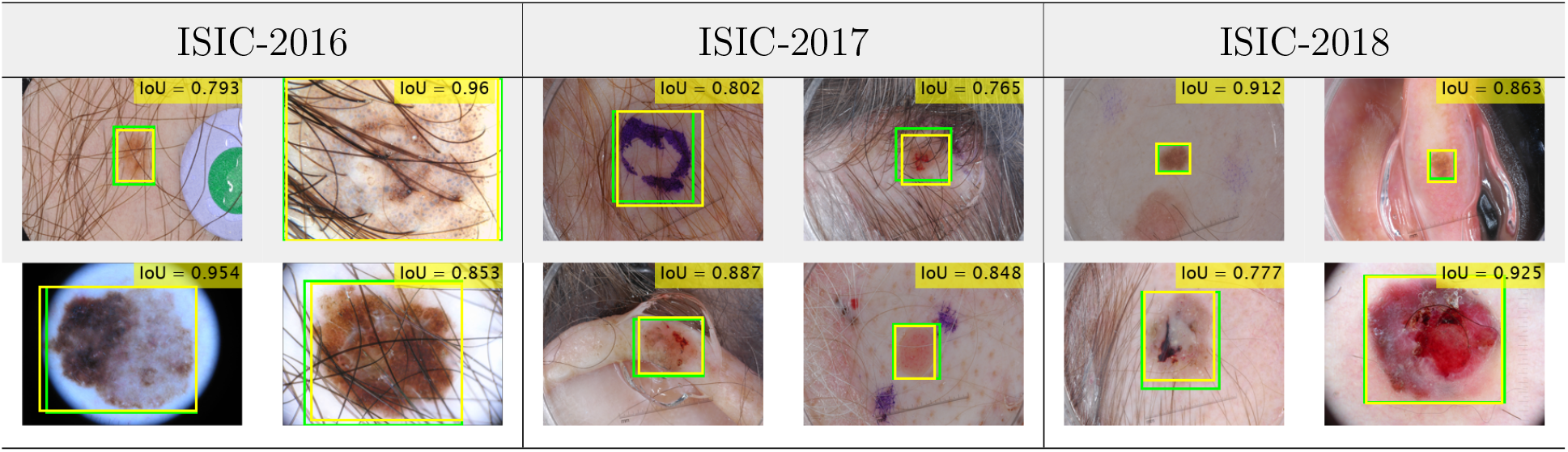
The example of some extracted ROIs on the ISIC-2016, ISIC-2017, and ISIC-2018 test datasets from the fine-tuned DSNet, where the green and yellow color denote the extracted and true bounding boxes, accordingly.

Fig. 10 bestows the representative examples of the augmented images of the segmented lesion ROIs. Those qualitative results reveal each image’s distinctiveness, which is crucial for the CNN training. Fig. 10 (d) shows that the sigmoid corrected image provides the specified lesion region to the network for learning about the lesion. The gamma-corrected image in Fig. 10 (f), for *γ* = 1.7, provides more intense lesion area, whereas Fig. 10 (e), for *γ* = 0.7, is brighter than the original image in Fig. 10 (a). The logarithmic corrected image (see in Fig. 10 (g)) is also darker, showing the distinctiveness.

**Figure 10:**
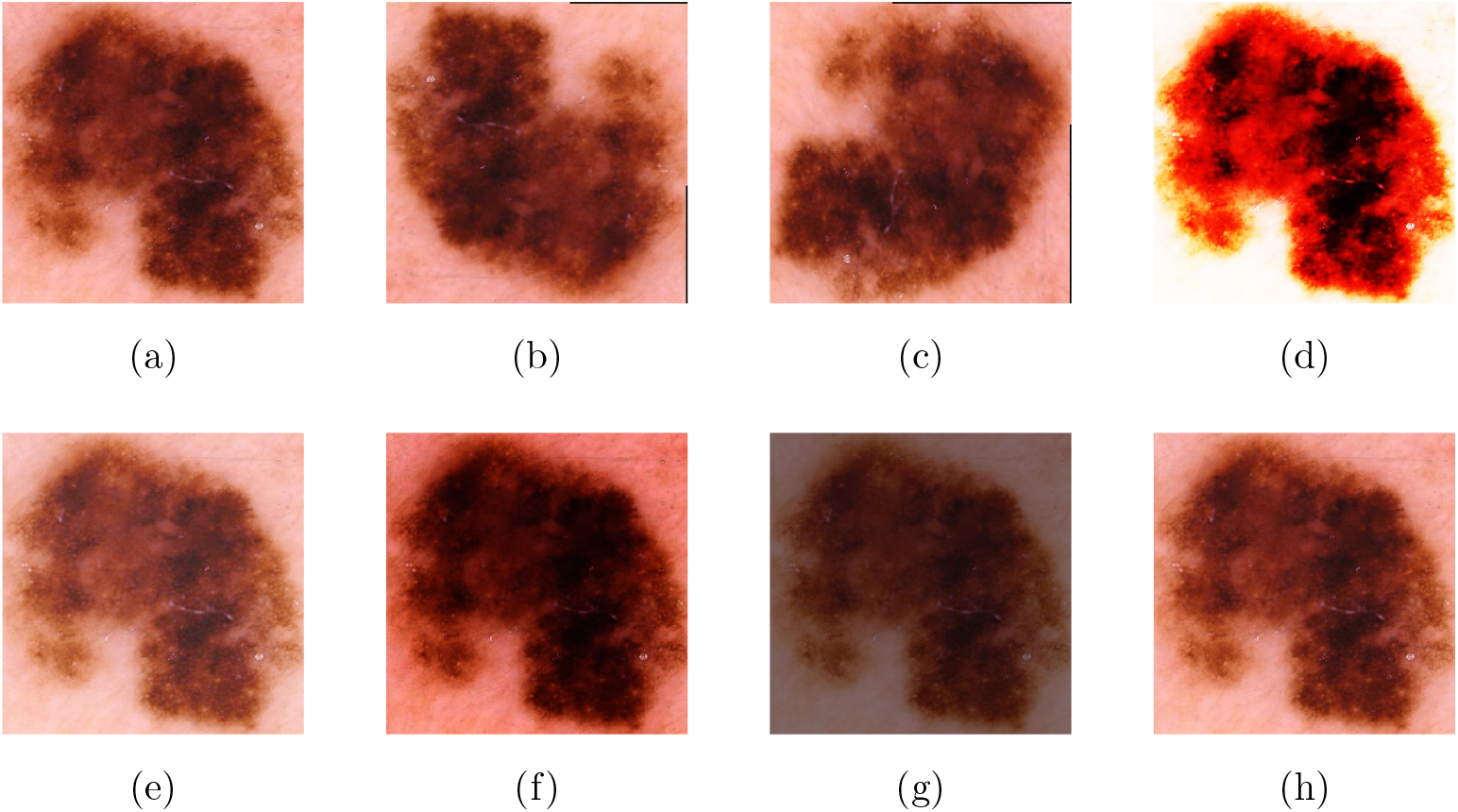
Examples of the geometry- and intensity-based augmentations of a dermoscopic image, where (a) the original image, (b) 180^*°*^ rotated, (c) 270^*°*^ rotated, (d) sigmoid corrected, (e) gamma corrected (*γ* = 0.7), (f) gamma corrected (*γ* = 1.7), (g) logarithmic corrected, and (h) intensity rescaled.

### 3.2. Optimizer and Learning Rate Selection

The Learning Rate (LR) is one of the essential hyperparameters, challenging to choose initially without experiments. Too small of an LR performs a slowly converging training, while too large of an LR makes the training algorithm diverge [54]. However, to update the weight parameters for minimizing the loss function, the optimizer is also very critical and crucial to select. We have done several experiments to get a better-LR and optimizer for the SLC. We have applied input (*I*_1_) and preprocessing (*P*_1_) to generate the output (*O*_1_) by employing different optimizers and LRs, as presented in Fig. 11. This experiment has been conducted using multiple optimizers, such as Stochastic Gradient Descent (SGD) [59], Adadelta [79], Adamax & Adam [36], and different LR scheduler schemes, such as constant LR, decaying LR with epochs, and Cyclical Learning Rates (CyLR) [65]. We set the initial epochs of 100 with the early stopping scheme for all the experiments. The training has been terminated when it stops improving the validation accuracy up to 10 epochs. The Adadelta optimizer, with an initial LR of 1.0, reaches the training accuracy of 100.0 % after few epochs, whereas the validation accuracy stagnates the improvement. Adam optimizer has also stopped improving training and validation accuracy with the increased epochs. The constant LR, decaying LR, and CyLR with SGD also produce many overshoots and undershoots in training and validation accuracies. On the other hand, the adaptive optimizer Adamax, with an initial LR of 0.0001, has smoothly increased training accuracy with the highest validation accuracy. We employ the LR scheduler (reduction of initial LR after 5 epochs if validation accuracy does not improve) along with the Adamax. However, those experiments reveal that an Adamax, with the LR scheduler, is a better choice for the SLC in our proposed DermoExpert, which is employed in the rest of the upcoming experiments.

**Figure 11:**
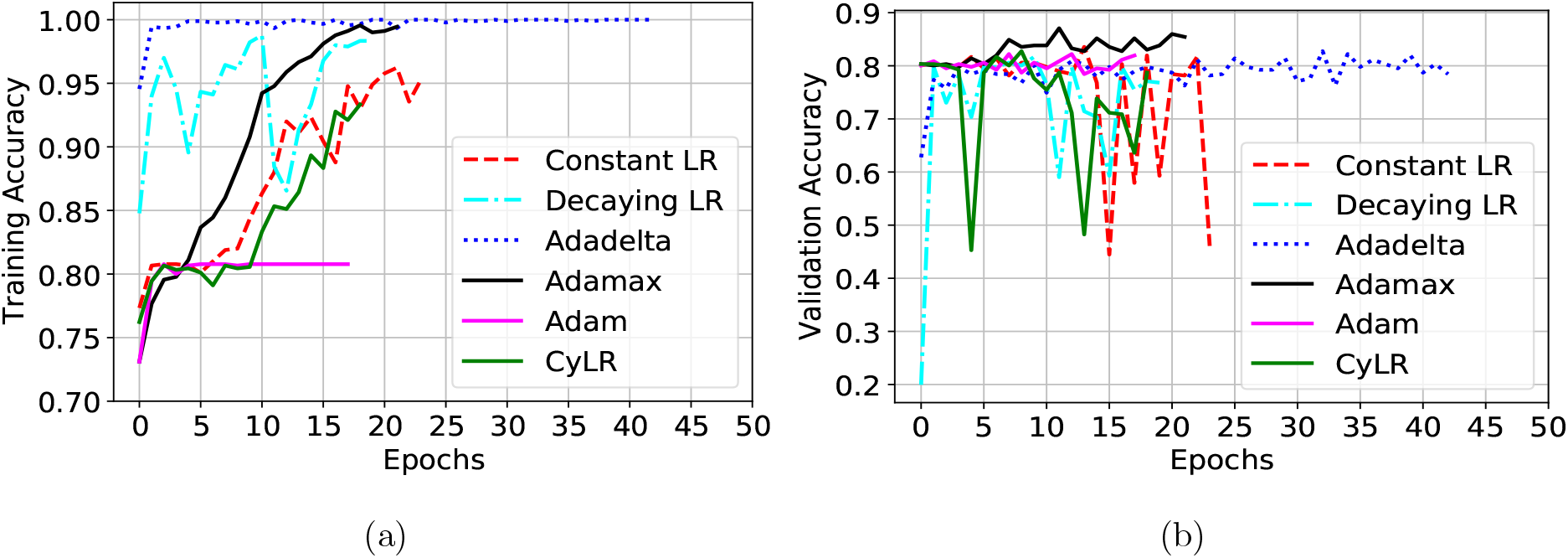
The experimental results for different optimizers and LR, where (a) is for the training accuracy, and (b) is for the validation accuracy.

#### 3.3. Classifier Selection

In this subsection, we exhibit the ablation studies on the proposed hybrid-CNN classifiers, as described in subsection 2.1.3, by comparing them quantitatively. We have used input (*I*_1_) and preprocessing (*P*_1_) to generate the output (*O*_1_) for five different classifiers, where we also apply the best optimizer and LR from the previous experiment. The results of this experiment are presented in the ROC curves in Fig. 12, where we have also announced AUCs for different classifiers. The results in Fig. 12 show that the Method-1 outperforms the Method-2 by a margin of 16.0 % in term of AUC. The true-positive rates are approximately 50.0 % and 25.0 % respectively for Method-1 and Method-2 for a 10.0 % false-positive rate (see in Fig. 12). Those results indicate that the channel-wise concatenation (*X*_*F C*_) is a better-fusion technique than the averaging (*X*_*F A*_) of the FMGs as the former has 3-times more depth information about the lesion. The comparison between the figures (see in Fig. 8 (b), Fig. 8 (c), Fig. 8 (d), Fig. 8 (e), and Fig. 8 (f)) also demonstrates that the addition of those feature maps produces a scattered feature distribution, whereas the individual feature map either from FMG-1, FMG-2, or FMG-3 depicts better-map of the lesion area (see in Fig. 8 (b)). Those phenomena could be the possible reasons for better results from the Method-1 than Method-2. Again, Method-3, where we have employed only the second-level ensembling, beats the former two methods, revealing that second-level ensembling has good prospects in the proposed classifier than the first-level ensembling alone. The employment of the fusion-by-averaging (*X*_*F A*_) with all three distinct feature maps, called Method-4, further improves Method-3 by a margin of 0.5 % concerning the AUC. Finally, in Method-5, if we replace *X*_*F A*_ by fusion-by-concatenation (*X*_*F C*_) in Method-4, it shows a better performance comparing all the former methods as *X*_*F C*_ has a better representation of the lesion than the *X*_*F A*_. However, the Method-5 beats all the other four methods by the margins of 8.3 %, 24.3 %, 7.0 %, and 6.5 % concerning AUC respectively for Method-1 to Method-4. The above discussions reveal that the hybrid-CNN classifier (Method-5) has a better-prospect for the SLC, comprising of both the first- and second-level ensembling. We have used fusion-by-concatenation in the first-level, and in the second-level, we aggregate the individual probability to get the final prediction probability. However, for all the next experiments for the SLC on the ISIC-2016, ISIC-2017, and ISIC-2018 datasets, we will use our proposed hybrid-CNN classifier (Method-5) as it has better-prospects as a classifier for the robust DermoExpert.

**Figure 12:**
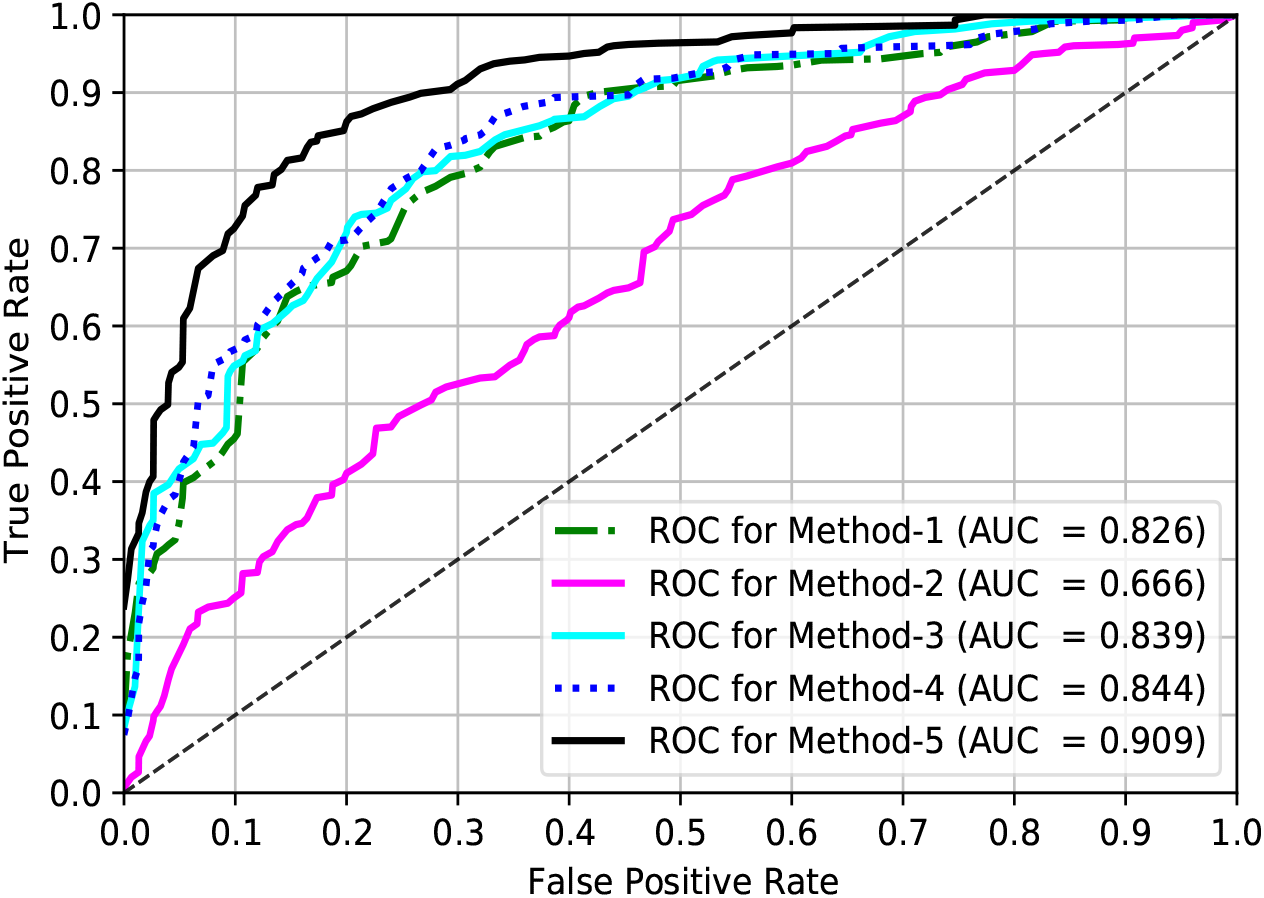
The ROC curves for the ISIC-2016 test dataset by employing the proposed five different networks and preprocessing (*P*_1_).

### 3.4. Results on ISIC-2016

The binary SLC’s performance in the proposed DermoExpert has been validated using 379 dermoscopic test images of the ISIC-2016 dataset. The overall quantitative results from all the extensive experiments are exhibited in Table 3 in terms of the recall, precision, and F1-score. The results in Table 3 show that the preprocessing (*P*_3_) along with the proposed terms of AUC. Also from Fig. 13 and given a 10.0 % false-positive rates, the true-positive rates of the proposed DermoExpert, Xception, ResNet-50, and DenseNet-121 are approximately 83.0 %, 55.0 %, 50.0 %, and 58.0 %, respectively. The above-discussions for the SLC on ISIC-2016 test dataset indicate that the proposed DermoExpert can be potentially used as an SLC-CAD tool for lesion classification.

**Table 3:** The classification results on the ISIC-2016 test dataset from the different extensive experiments.

| Preprocessing | Recall |  |  | Precision |  |  | F1-score |  |  |
| --- | --- | --- | --- | --- | --- | --- | --- | --- | --- |
|  | Nev | Mel | W. Avg. | Nev | Mel | W. Avg. | Nev | Mel | W. Avg. |
| $P_1$ | 0.99 | 0.35 | 0.87 | 0.86 | 0.93 | 0.87 | 0.92 | 0.50 | 0.84 |
| $P_2$ | 0.98 | 0.69 | 0.93 | 0.93 | 0.91 | 0.93 | 0.96 | 0.79 | 0.92 |
| $P_3$ | 0.93 | 0.85 | 0.92 | 0.96 | 0.76 | 0.92 | 0.95 | 0.81 | 0.92 |

$P_1$ : Segmentation; $P_2$ : Segmentation and Rebalancing; $P_3$ : Segmentation, Rebalancing, and Augmentation.

**Figure 13:**
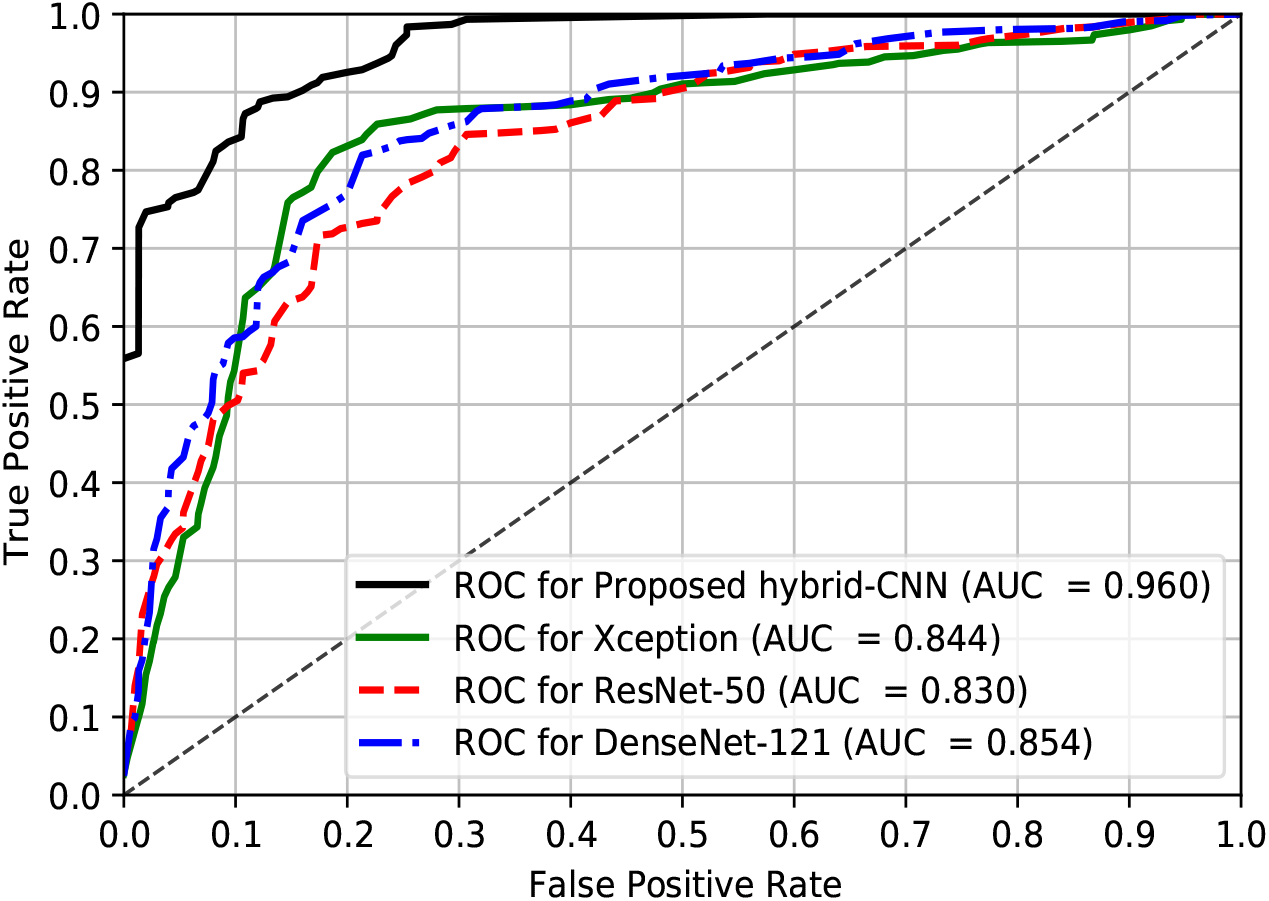
The ROC curves for the ISIC-2016 test dataset by employing the proposed CNN-based hybrid classifier and preprocessing (*P*_3_).

### 3.5. Results on ISIC-2017

This subsection represents the potentiality of the proposed DermoExpert to recognize three different lesions as Nev, SK, and Mel. The quantitative results on the ISIC-2017 test dataset have been summarized in Table 5 using different metrics, such as recall, precision, and F1-score. As in Table 5, the results depict the weighted average metrics for aggregate cases concerning the class population. The SLC results, as shown in Table 5, demonstrate that the hybrid-CNN classifier yields the best performance for the binary SLC. The recall of the positive class (Mel) reveals that the type-II errors are 65.0 %, 31.0 %, and 15.0 % for the respective preprocessing *P*_1_, *P*_2_, and *P*_3_. It also points that rebalancing with segmentation improves the type-II errors by 34.0 %, whereas the rebalancing and augmentation with segmentation significantly improve the type-II errors by 50.0 %. Although the FN reduces when we move from *P*_1_ to *P*_3_, the FP increases accordingly. For a *P*_3_ preprocessing, the Mel class’s precision (76.0 %) also shows the evidence of increasing FP with the decreasing of FN, as 24.0 % recognized positives are the wrong positives. However, the decreasing FN-rates (65.0 % to 15.0 %) is better than the increasing FP-rates (1.0 % to 7.0 %) in medical diagnosis applications. Moreover, the harmonic mean of the precision and recall for both the classes (Nev and Mel) is improving when we move from *P*_1_ to *P*_3_ by the margins of 3.0 % and 31.0 % respectively for Nev and Mel classes. The confusion matrix in Table 4, for more detailed analysis of the SLC-2016 results, shows that among 304 Nev samples, correctly classified samples are 284 (93.4 %), whereas only 20 (6.58 %) samples are incorrectly classified as Mel. It also shows that among 75 Mel samples, rightly classified samples are 64 (85.33 %), whereas only 11 (14.67 %) samples are mistakenly classified as Nev. Fig. 13 shows the ROC curves of the best SLC-2016 and the baseline Xception [9], ResNet-50 [25], and DenseNet-121 [29]. The proposed DermoExpert obtains an AUC of 0.96, indicating the probability of correct lesion recognition is as high as 96.0 % for any given random sample. It has beaten the baseline Xception, ResNet-50, and DenseNet-121 respectively by 11.6 %, 13.0 %, and 10.6 % in recall is increased by 24.0 %, and 17.0 % respectively for Mel and SK due to the employment of preprocess *P*_2_ instead of baseline *P*_1_. The further employment of the preprocess *P*_3_ in place of *P*_2_ could not reduce the type-II errors somewhat remains constant, but reduces type-II errors of the Nev class by a border of 5.0 %. The weighted average recall has been increased when we employ the preprocess *P*_2_ instead of baseline *P*_1_, and then the preprocess *P*_3_ instead of baseline *P*_2_. However, it is beneficial to apply the preprocess *P*_3_ instead of *P*_2_ and *P*_1_ in terms of type-II errors. It is noticed from Table 5 that the weighted precisions are increasing, when we change the preprocess *P*_1_ to *P*_2_ and *P*_2_ to *P*_3_. The class-wise precision also reveals that the FN is reducing significantly, although the FP is increasing. However, such a result in medical disease diagnosis for the SLC is acceptable as the patients with the positive symptom should not be classified as negative patients. Additionally, the class-wise F1-score for the Nev, SK, and Mel have improved significantly, when we change the prepossess *P*_1_ to *P*_2_, and then *P*_2_ to *P*_3_. The improved F1-score tells that both the recall and precision are praiseworthy, although the uneven class distribution was being used. Details of class-wise investigation of the best performing SLC has been present in the confusion matrix in Table 6, applying the preprocessing (*P*_3_) and the proposed hybrid-CNN classifier. The confusion matrix in Table 6 for the SLC-2017 shows that 94.66 % Nev samples are correctly classified as Nev, whereas 5.34 % (3.31 % as SK and 2.03 % as Mel) samples are wrongly classified. 84.44 % SK samples are correctly categorized as SK, whereas 15.56 % (6.67 % as Nev and 8.89 % as Mel) samples are improperly classified. On the other hand, 60.69 % Mel samples are genuinely classified as Mel, whereas 39.31 % (29.91 % as Nev and 9.40 % as SK) samples are awkwardly classified. Although the 39.31 % of the positive samples (Mel) are wrongfully classified, it is still better than the baseline 62.0 % errors in the baseline preprocessing *P*_1_. Fig. 14 shows the ROC curves of the best SLC-2017 and baseline state-of-the-art Xception, ResNet-50, and DenseNet-121. The proposed DermoExpert for the SLC-2017 obtains an AUC of 0.947, designating the probability of correct lesion recognition is as high as 94.7 % for any given random sample. The proposed DermoExpert outperforms the Xception, ResNet-50, and DenseNet-121 respectively by 10.0 %, 12.3 %, and 19.7 % for AUC. Also from Fig. 14 and given a 10.0 % false-positive rate, the true-positive rates of the proposed DermoExpert, Xception, ResNet-50, and DenseNet-121 are approximately 85.0 %, 60.0 %, 55.0 %, and 41.0 %, respectively. In contrast, the above-discussions for the SLC on ISIC-2017 demonstrate that the proposed DermoExpert can be potentially applied as an SLC-CAD tool.

**Table 4:** The confusion matrix for the ISIC-2016 test dataset by using the proposed CNN-based hybrid classifier and preprocessing (*P*_3_).

|  |  | Actual |  |
| --- | --- | --- | --- |
|  |  | Nev | Mel |
| Predicted | Nev | 284<br>93.42 % | 11<br>14.67 % |
|  | Mel | 20<br>6.58 % | 64<br>85.33 % |

**Table 5:**
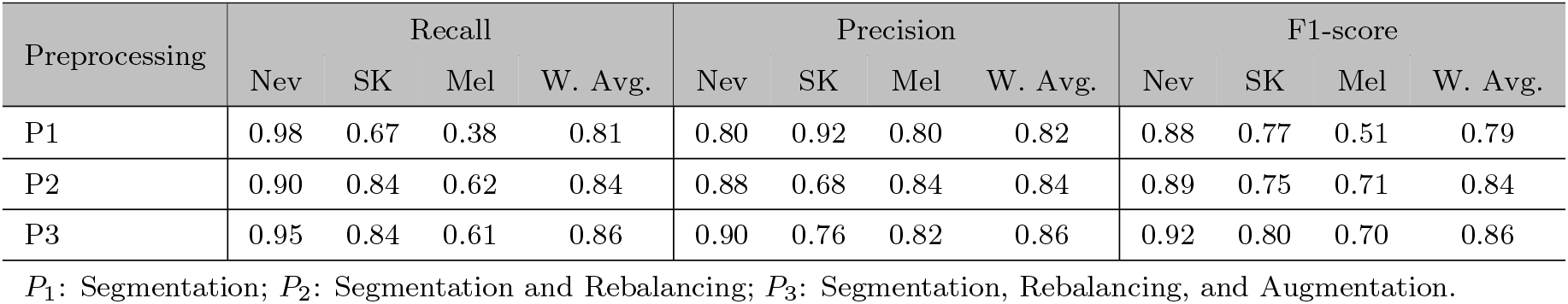
The classification results on the ISIC-2017 test dataset from the different extensive experiments.

| Preprocessing | Recall |  |  |  | Precision |  |  |  | F1-score |  |  |  |
| --- | --- | --- | --- | --- | --- | --- | --- | --- | --- | --- | --- | --- |
|  | Nev | SK | Mel | W. Avg. | Nev | SK | Mel | W. Avg. | Nev | SK | Mel | W. Avg. |
| P1 | 0.98 | 0.67 | 0.38 | 0.81 | 0.80 | 0.92 | 0.80 | 0.82 | 0.88 | 0.77 | 0.51 | 0.79 |
| P2 | 0.90 | 0.84 | 0.62 | 0.84 | 0.88 | 0.68 | 0.84 | 0.84 | 0.89 | 0.75 | 0.71 | 0.84 |
| P3 | 0.95 | 0.84 | 0.61 | 0.86 | 0.90 | 0.76 | 0.82 | 0.86 | 0.92 | 0.80 | 0.70 | 0.86 |

$P_1$ : Segmentation; $P_2$ : Segmentation and Rebalancing; $P_3$ : Segmentation, Rebalancing, and Augmentation.

**Table 6:** The confusion matrix for the ISIC-2017 test dataset by using the proposed hybrid-CNN classifier and preprocessing (*P*_3_).

|  |  | Actual |  |  |
| --- | --- | --- | --- | --- |
|  |  | Nev | SK | Mel |
| Predicted | Nev | 372 | 6 | 35 |
|  |  | 94.66 % | 6.67 % | 29.91 % |
|  | SK | 13 | 76 | 11 |
|  |  | 3.31 % | 84.44 % | 9.40 % |
|  | Mel | 8 | 8 | 71 |
|  |  | 2.03 % | 8.89 % | 60.69 % |

**Figure 14:**
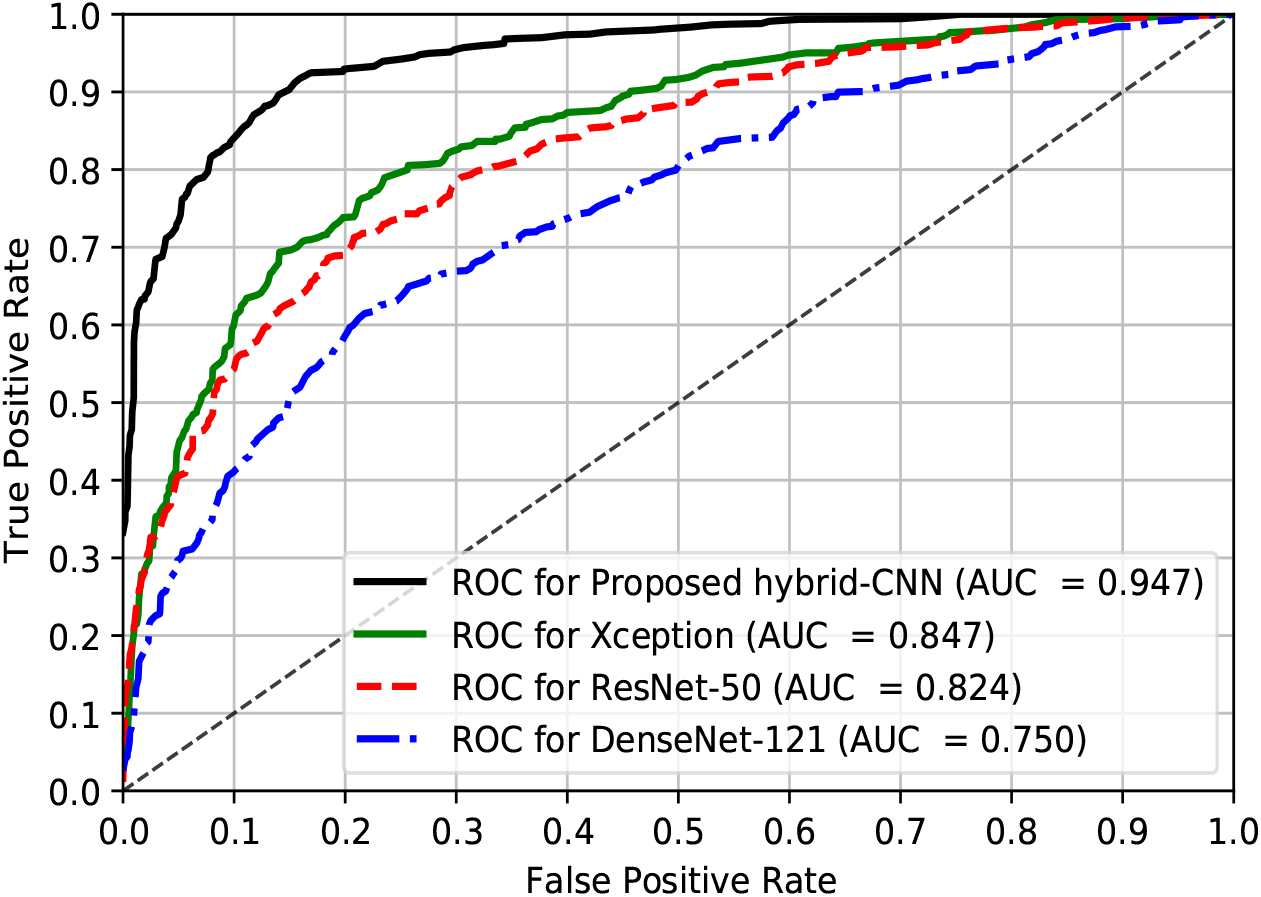
The ROC curves for the ISIC-2017 test dataset by employing the proposed classifier and preprocessing (*P*_3_).

### 3.6 Results on ISIC-2018

This subsection represents the experimental results for the SLC-2018 from the proposed DermoExpert for recognizing very challenging lesion categories into seven classes, such as Nev, SK, BCC, AK, DF, VL, and Mel. As presented earlier, in subsection 2.1.1, the ISIC-2018 provides only the training set. Therefore, we employed 5-fold cross-validation, where 60.0 %, 20.0 %, and 20.0 % samples are utilized respectively for training, validation, and testing. We repeat the experiments 5-times for the SLC-2018, and the final results in Fig. 15 are the average classification result of the 5-folds. The investigation on the results (see in Fig. 15) explicates that for a baseline preprocessing (*P*_1_), the metrics (see blue bar in Fig. 15 (a), Fig. 15 (b), and Fig. 15 (c)) are varied in significant margins, where the recall, precision, and F1-score of the Nev class are higher than the other classes. Moreover, the melanoma class has a recall of 42.0 %, which is significantly less. Additionally, the recall, precision, and F1-score for the DF class are 0.0 %, where all the DF class images are classified as other classes. Such a weak result from the proposed DermoExpert by applying the baseline preprocessing (*P*_1_) is due to the unequal class distribution, where class distribution was 1.0 : 6.1 : 13.02 : 20.47 : 58.3 : 47.47 : 6.02 respectively for the classes Nev, SK, BCC, AK, DF, VL, and Mel. In terms of recall, precision, and F1-score, the DF and VL classes’ weak performance is likely to happen as they are the most minority (underrepresented) classes. However, the rebalancing of the class distribution, as in the preprocessing (*P*_2_), boosts the class-wise performance in significant portions (see green bar in Fig. 15 (a), Fig. 15 (b), and Fig. 15 (c)), where the recall, precision, and F1-score of the DF class respectively increase to 65.0 %, 64.0 %, and 64.0 % from the baseline 0.0 % for all metrics. Not only the DF class but also the other classes have improved the performance, especially the recalls of BCC, AK, and Mel class have increased by the margins of 33.0 %, 29.0 %, and 11.0 %, respectively. Another point can be claimed that moving the preprocessing, from *P*_1_ to *P*_2_, does not degrade the best performing Nev class’s performance in *P*_1_. Instead, it improves in terms of precision and F1-score, while the recall remains constant. However, the proposed augmentations with the preprocessing (*P*_2_) improve all the metrics for most classes, while the other classes’ performance remain constant. The more enhanced class-wise precision and F1-score, due to the third preprocessing appliance (*P*_3_), confirm that the positive predictive value and the balanced precision-recall are also increased than the type-II errors (recall). The more detailed class-wise assessment of the best performing SLC for the ISIC-2018 test dataset has been presented in the confusion matrix in Table 7, employing the preprocessing (*P*_3_) and the proposed hybrid-CNN classifier. The matrix in Table 7 reveals the FN and FP for the SLC-2018, where number of wrongly classified images (type-I or type-II errors) are 58*/*1341 (4.33 %), 72*/*220 (32.73 %), 18*/*102 (17.65 %), 29*/*65 (44.62 %), 7*/*23 (30.43 %), 1*/*29 (3.45 %), and 102*/*222 (45.94 %) respectively for the Nev, SK, BCC, AK, DF, VL and Mel. Although some classes’ performance is not highly improved as in other classes, it is still better than the baseline process. Fig. 16 shows the ROC curves of the best SLC-2018 and baseline state-of-the-art Xception, ResNet-50, and DenseNet-121. The proposed DermoExpert for the SLC-2018 achieves an AUC of 0.969, which designates the probability of correct lesion recognition is as high as 96.9 % for any given random sample. It has defeated all the baseline Xception, ResNet-50, and DenseNet-121 respectively by 2.4 %, 2.4 %, and 2.3 % with respect to AUC. Also from Fig. 16 and given a 10 % false-positive rate, the true-positive rates of the proposed DermoExpert, Xception, ResNet-50, and DenseNet-121 are approximately 92.0 %, 83.0 %, 82.0 %, and 86.0 %, respectively. In conclusion, the above-discussions for the SLC on ISIC-2018 demonstrate that the proposed DermoExpert can be potentially applied as an SLC-CAD tool.

**Table 7:** The confusion matrix for the ISIC-2018 test dataset by using the proposed classifier and prepro-cessing (*P*_3_).

|  |  | Actual |  |  |  |  |  |  |
| --- | --- | --- | --- | --- | --- | --- | --- | --- |
|  |  | Nev | SK | BCC | AK | DF | VL | Mel |
| Predicted | Nev | 1283 | 43 | 10 | 3 | 4 | 1 | 64 |
|  |  | 95.67 % | 19.55 % | 9.80 % | 4.62 % | 17.39 % | 3.45 % | 28.83 % |
|  | SK | 21 | 148 | 7 | 11 | 2 | 0 | 27 |
|  |  | 1.57 % | 67.27 % | 6.86 % | 16.92 % | 8.69 % | 0.0 % | 12.16 % |
|  | BCC | 9 | 4 | 84 | 8 | 0 | 0 | 3 |
|  |  | 0.67 % | 1.82 % | 82.36 % | 12.31 % | 0.0 % | 0.0 % | 1.35 % |
|  | AK | 0 | 9 | 0 | 36 | 0 | 0 | 3 |
|  |  | 0.0 % | 4.09 % | 0.0 % | 55.38 % | 0.0 % | 0.0 % | 1.35 % |
|  | DF | 5 | 2 | 0 | 1 | 16 | 0 | 0 |
|  |  | 0.37 % | 0.91 % | 0.0 % | 1.54 % | 69.57 % | 0.0 % | 0.0 % |
|  | VL | 0 | 0 | 0 | 0 | 0 | 28 | 5 |
|  |  | 0.0 % | 0.0 % | 0.0 % | 0.0 % | 0.0 % | 96.55 % | 2.25 % |
|  | Mel | 23 | 14 | 1 | 6 | 1 | 0 | 120 |
|  |  | 1.72 % | 6.36 % | 0.98 % | 9.23 % | 4.35 % | 0.0 % | 54.06 % |

**Figure 15:**
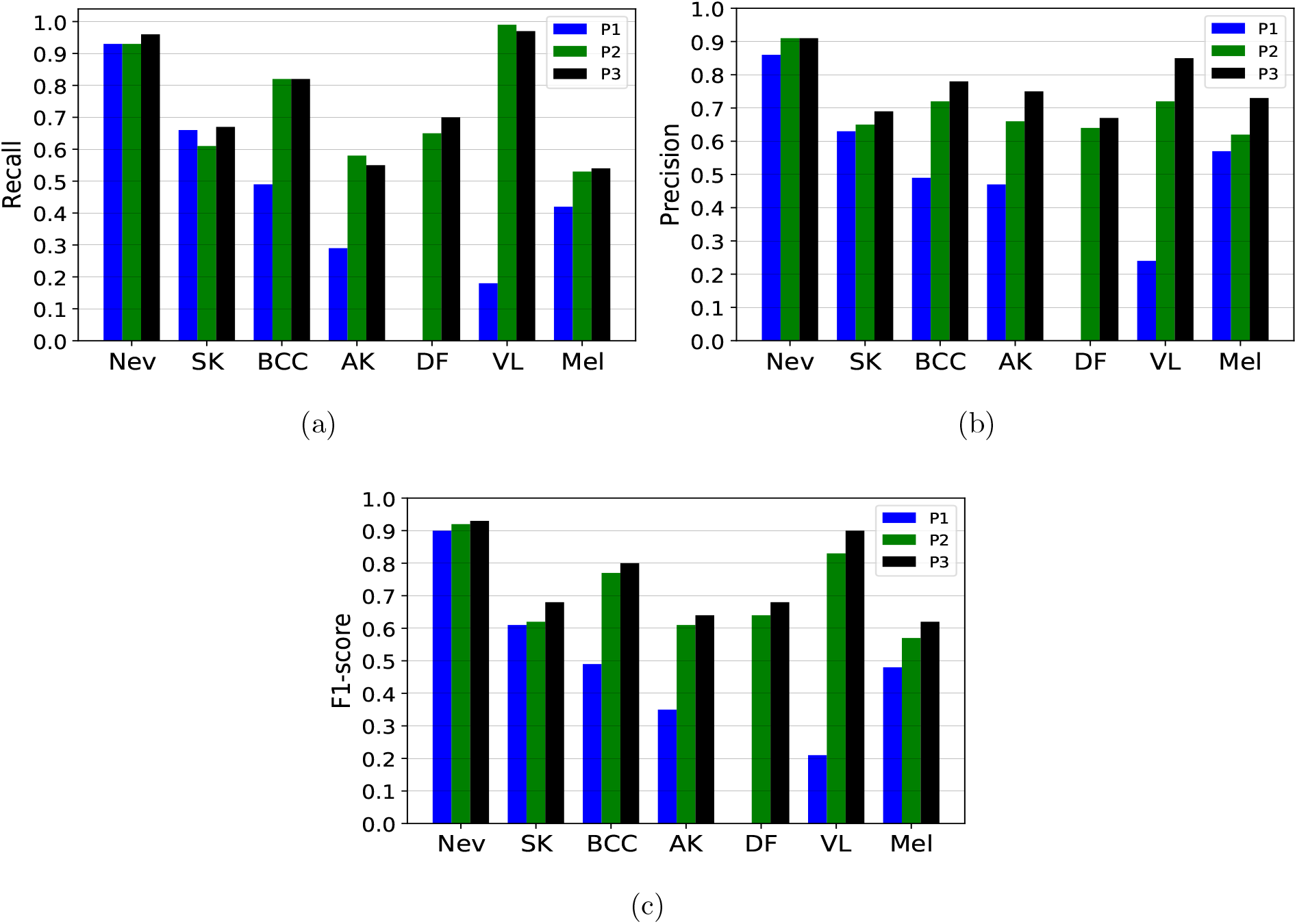
The experimental results for the ISIC-2018 test dataset classification, where blue, green, and black bar of different class respectively denote the results (recall (a), precision (b), and F1-score (c)) for the preprocessing *P*_1_, *P*_2_, and *P*_3_.

**Figure 16:**
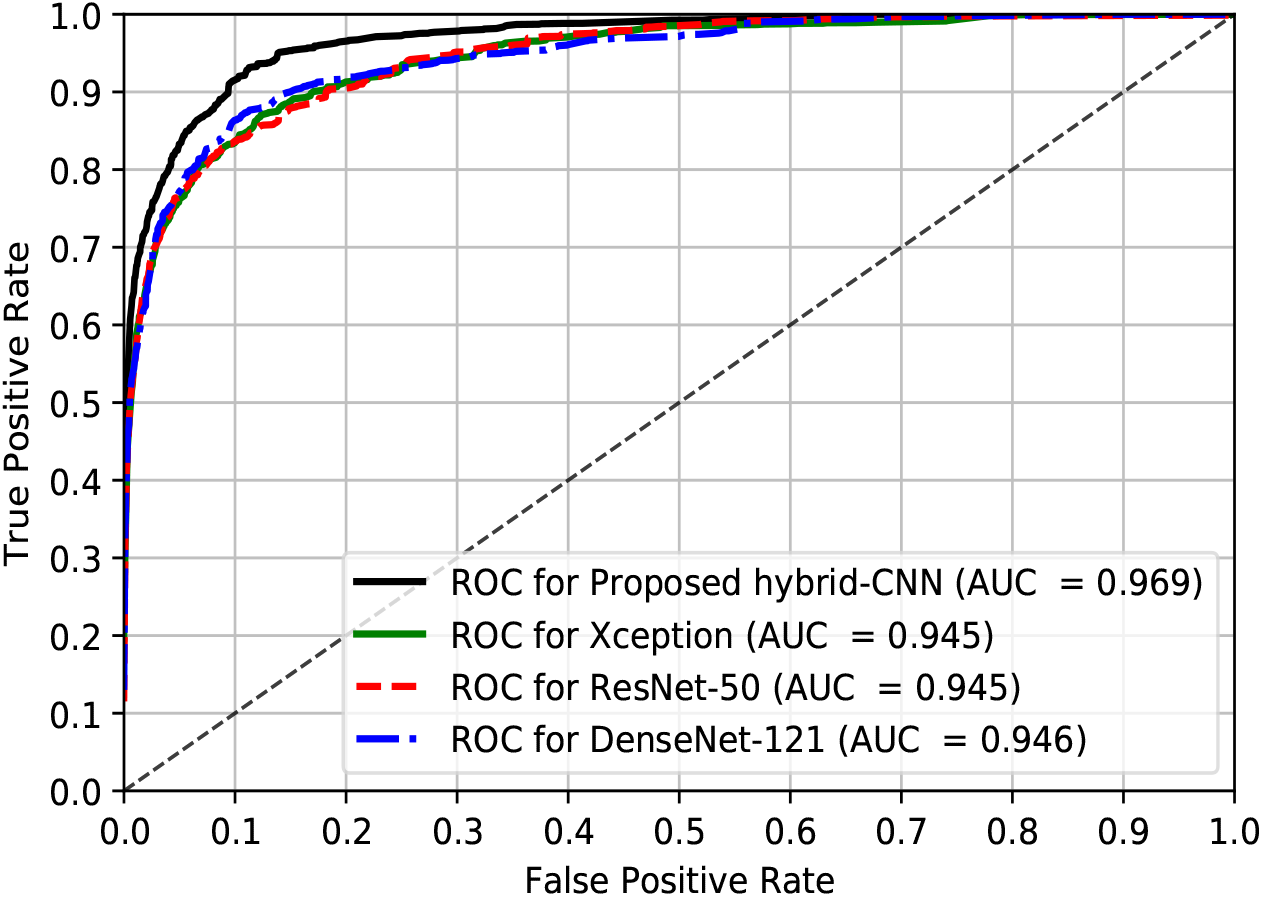
The ROC curves for the ISIC-2018 test dataset by employing the proposed network and prepro-cessing (*P*_3_).

### 3.7. Results Comparison

In this subsection, the performance of the proposed DermoExpert is compared and contrasted to several current state-of-the-art methods.

Table 8 represents the performance comparison of the proposed DermoExpert with other recent methods for all the ISIC-2016, ISIC-2017, and ISIC-2018 datasets. To improve classification performance, authors in several recent works used the external data to train their models, which are not publicly available yet. The improvement of the classification network may not be due to the network’s superiority, but the external data characteristics, which are similar to the test datasets. However, for fairness in comparison, we have reported the results, which were too strict for the datasets on the ISIC archive only. The proposed DermoExpert produces the best classification, as shown in Table 8, for five out of the nine cases while performing second best with the winning methods on the other four cases.

**Table 8:** The state-of-the-art comparison with proposed DermoExpert, which were trained, validated and tested on the ISIC-2016, ISIC-2017, and ISIC-2018 datasets.

| Methods | ISIC-2016 |  |  | ISIC-2017 |  |  | ISIC-2018 |  |  |
| --- | --- | --- | --- | --- | --- | --- | --- | --- | --- |
|  | Recall | Precision | AUC | Recall | Precision | AUC | Recall | Precision | AUC |
| ResNet-50 [7] | 0.56 | 0.71 | 0.85 | - | - | - | - | - | - |
| GR [60] | - | - | - | 0.15 | - | 0.91 | - | - | - |
| ARLCNN [80] | - | - | - | 0.77 | - | 0.92 | - | - | - |
| IR [2] | 0.82 | - | 0.77 | 0.76 | - | - | 0.81 | - | - |
| FPRPN [66] | **0.99** | <u>0.82</u> | 0.81 | **0.98** | <u>0.82</u> | 0.79 | - | - | - |
| GCD [70] | 0.32 | 0.73 | <u>0.86</u> | 0.55 | - | <u>0.93</u> | - | - | - |
| MFA [78] | 0.60 | 0.69 | <u>0.86</u> | - | 0.72 | 0.90 | - | - | - |
| MIAG [49] | - | - | - | - | - | - | 0.78 | **0.87** | **0.98** |
| DenseNet [40] | - | - | - | - | - | - | 0.80 | <u>0.85</u> | **0.98** |
| RDI [53] | - | - | - | - | - | - | <u>0.83</u> | 0.83 | **0.98** |
| **DermoExpert (Proposed, 2020)** | <u>0.92</u> | **0.92** | **0.96** | <u>0.86</u> | **0.86** | **0.95** | **0.86** | <u>0.85</u> | <u>0.97</u> |

**GR:** Gabor Wavelet-based CNN [60]
**ARLCNN:** Attention residual learning convolutional neural network (ResNet-14 & ResNet-50) [81]
**IR:** Inception-ResNet-V2 (ISIC-2016), ResNet-50 (ISIC-2017,ISIC-2018) [2]
**FPRPN:** Feature Pyramid Network (FPN) and Region Proposal Network (RPN) [66]
**GCD:** Global-Part CNN Model with Data-Transformed Ensemble Learning [70]
**MFA:** Multi-network based feature aggregation [78]
**MIAG:** Emsembling classifier [49]
**RDI:** Emsembling of ResNet, DenseNet, and Inception [53]

#### Comparison of SLC-2016

The proposed DermoExpert produces the best results for the AUC by beating the state-of-the-art [70, 78] with a 10.0 % margin. Concerning the type-II errors (recall), DermoExpert is behind the state-of-the-art [66] by 7.0 %, but the DermoExpert outperforms the FPRPN [66] by a 10.0 % margin concerning the positive predictive value (precision). However, in terms of balanced accuracy (avg. of recall and precision), DermoExpert beats the state-of-the-art [66] by a 1.5 % margin.

#### Comparison of SLC-2017

For AUC, the DermoExpert wins by defeating the second-highest [70] by a margin of 2.0 %. DermoExpert has beaten the work of Song et al. [66] by the margins of 4.0 %, and 16.0 % with respective precision and AUC although it lost by Song et al. [66] in terms of recall. However, the proposed DermoExpert produces the second-highest results by beating the third results [80] with a margin of 9.0 % for recall.

#### Comparison of SLC-2018

The results of DermoExpert for ISIC-2018 are compared with the top three performers of the ISIC-2018 competition leaderboard [40, 49, 53], where DermoExpert wins in recall and serves as a second winner in the other two metrics. It also beats the recently published method of Al-Masni et al. [2] by a margin of 5.0 % in terms of type-II errors. It means that 5.0 % additional samples will be classified correctly as the actual class than the method of Al-Masni et al. [2]. However, in terms of the balanced accuracy, DermoExpert beats the challenge topper by a margin of 3.0 %. The recent method proposed by Mahbod et al. [46], for the test results of ISIC-2018, are not presented in Table 8, as they did not use the same dataset to train their model, as in the proposed DermoExpert.

## 4. Discussion and Application

CNN-based classifiers are better-choice in different medical imaging context, where they automatically learn low-, middle-, and high-level features directly from the input images. Finally, fully connected neural networks, also known as multilayer perceptron, classify those features. However, such deep CNN-based classifiers’ training is an arduously challenging process, especially when training with a smaller dataset, such as the ISIC skin lesion datasets. There are two commonly occurring limitations in the CNN-based classifiers: prone to over-fitting and vanishing gradient problems. Those two critical limitations are reduced for the SLC in this article.

We proposed a hybrid classifier to build a generic and robust end-to-end SLC system utilizing a smaller dataset. We use three distinct feature map-generators rather than a single generator as in Xception, ResNet-50, DenseNet-121, and *etc*. In each generator in the proposed classifier, we employ several skip connections, enabling each layer of the generator to access the gradients of all previous layers directly. However, such a CNN network design is more likely to alleviate the vanishing gradient problem and also partly reduce the overfitting as the final feature map has a more in-depth skin lesion presentation. The adaptation of pre-trained weights in place of the CNN classifiers training from scratch, with any random initialization, also partially reduces the overfitting and improves the performance. As in subsection 3.3, the experimental results have validated that multiple generators’ multiple features provide better results. Moreover, channel-wise concatenation of different features is better than the addition of them as the latter one generates scattered feature maps (see in Fig. 8). In the same experimental conditions, the former approach outperforms the latter approach by a margin of 16.0 % concerning AUC (see in Fig. 12). Further addition of second-level ensembling, as described in subsection 2.1.3, provides 7.0 % more AUC than first- or second-level ensembling alone (see in Fig. 12). Instead of a random selection of LR and optimizer, it is better to conduct several extensive experiments. The cost functions depend on the data distribution, size, and inter- or intra-class variability. However, our experimental results, as presented in subsection 3.2, have demonstrated that the adaptive optimizer Adamax, with an initial LR of 0.0001, has a better-prospect for the SLC (see in Fig. 11) when the LR scheduler is incorporated with the Adamax.

The classification results by the proposed DermoExpert, for the ISIC-2016 (see in subsection 3.4), ISIC-2017 (see in subsection 3.5), and ISIC-2018 (see in subsection 3.6), confirm that class rebalancing along with the segmentation enhances the performance of underrepresented class. The F1-score of the test datasets of ISIC-2016, ISIC-2017, and ISIC-2018 are improved by the margins of 8.0%, 5.0 %, and 1.0 %, respectively, when we have united additional images to underrepresented class and weighted the loss function. Further addition of image augmentation, with segmentation and rebalancing, improves the classification performance. It also reduces the overfitting, for the lesion classification, by reducing the differences between the training and testing performances. For all the test datasets of lesion classification, the proposed DermoExpert, with multiple feature map generators and two-level ensembling, performs better than the classifier having a single generator (see in Fig. 13, Fig. 14, and Fig. 16). The experimental results, in subsections 3.4, 3.5, and 3.6, also demonstrate that the classification performance for ISIC-2016 (two classes) is better than ISIC-2017 (three classes). The addition of SK class, as in ISIC-2017, reduces the F1-score of Nev and Mel classes respectively by the margins of 3.0 %, and 11.0 %. The higher similarity of SK with Nev and Mel classes is causing such a reduction in ISIC-2017 test results. Moreover, many classes tend to bring complications in the classifiers, especially when training with fewer examples and intra-class similarity, as in the SLC on the ISIC skin lesion datasets. It is also observable that the classification performance for ISIC-2018 has improved, although it has more classes (7 classes). However, the distribution of all the ISIC datasets (see in Table 1) depicts that the ISIC-2018 has much higher samples than the ISIC-2016 and ISIC-2017 datasets.

However, those discussions reveal the superiority of the proposed DermoExpert for the SLC, showing its acceptance for the SLC-CAD system. A few qualitative results of the proposed DermoExpert are illustrated in Fig. 17, where the segmented masks from the fine-tuned DSNet are used to detect the lesion ROI (yellow color contour) along with the recognized class. More classification results for all the test images are available on YouTube (ISIC-2016^5^, ISIC-2017^6^, and ISIC-2018^7^). Fig. 17 also shows a few challenging images to be classified and some wrongly recognized images. Although the DermoExpert wrongly predicts those images, they visually seem like the predicted class. However, in this article, we have presented the future applications of the DermoExpert by building a web application, as shown in Fig. 18 (a), for the user of the DermoExpert. We have implemented the app in our local machine, which runs in a web browser at “http://127.0.0.1:5000/” by accessing the CNN environments of that local machine. Our source code and segmented masks of the ISIC-2018 dataset will be publicly available in GitHub^8^.

**Figure 17:**
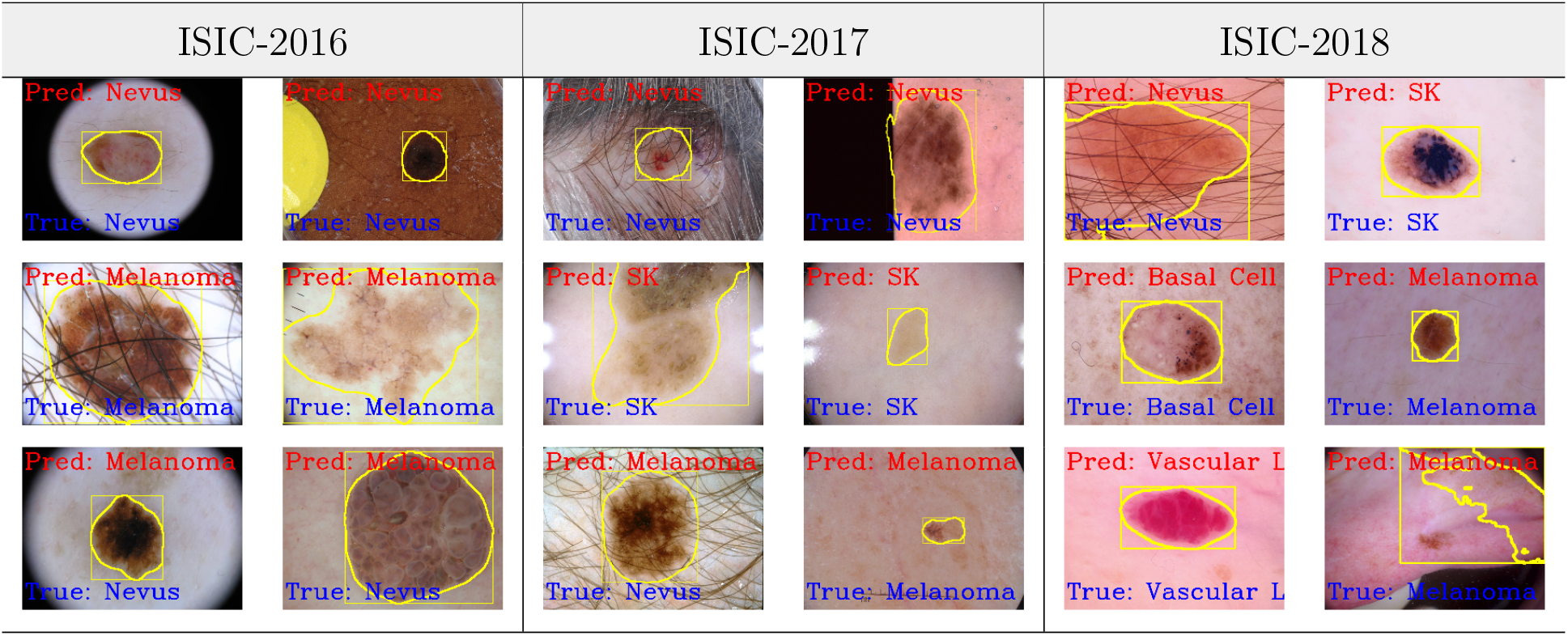
The qualitative classification results using the DermoExpert, where the classification has been accomplished using the segmented ROIs (yellow color) and the proposed DermoExpert.

**Figure 18:**
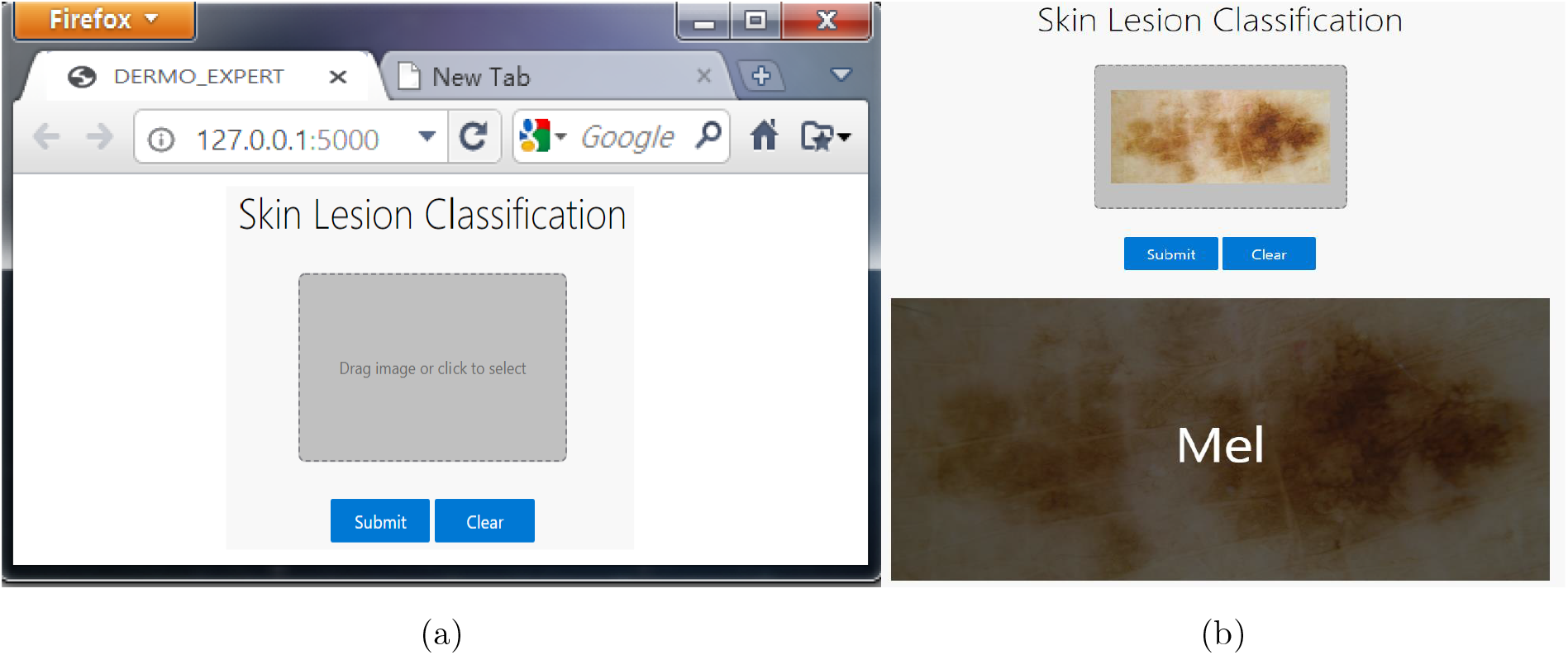
The web app for predicting the lesion class by employing the trained weights of the DermoExpert. Users can drag or choose an image by clicking the button, as shown on the left side, and can clear if the selection is wrong. After that, by clicking the submit button, the user can see the recognized result on the screen, as shown on the right side.

## 5. Conclusion

Although it is very challenging due to high visual similarity and diverse artifacts, skin lesions’ automatic recognition is significant. However, in this article, the skin lesion recognition has been automated by proposing a pipeline called DermoExpert. This article emphasized a systematic evaluation of an integrated skin lesion recognition system, including lesion ROI extraction, image augmentation, rebalancing, and hybrid-CNN classifier. Our experimental results demonstrate that the proposed DermoExpert can discriminate against the lesion features more accurately as we concatenated features from three distinct generators. Thus, it achieves state-of-the-art performance for the SLC of three different datasets. The segmented skin lesions, rather than the whole images, can provide more salient and representative features from the CNNs, leading to the SLC’s improvement. Moreover, the rebalanced class distribution attained better SLC performance compared to the imbalanced distribution. Additionally, the augmentation can lead the CNN-based classifier to be more generic as CNNs can learn from diverse training samples. We will further explore and investigate the effects of improving segmentation and weighting of the underrepresented classes in the future. The weights of the DermoExpert will be deployed to the Google Cloud platform to make it publicly available. We will provide our segmented masks of the ISIC-2018 dataset (Task 3: Lesion Diagnosis (10015 images)) for the research purpose (on-request) as they are not available yet. The proposed framework will be applied to other domains for medical imaging to verify its versatility and generality.

## Data Availability

None

## CRediT authorship contribution statement

**M. K. Hasan:** Conceptualization, Methodology, Software, Formal analysis, Investigation, Visualization, Writing-Review & Editing, Supervision; **M. T. E. Elahi:** Validation, Data Curation, Writing-Original Draft; **M. A. Alam:** Validation, Data Curation, Writing-Original Draft; **M. T. Jawad:** Writing-Original Draft, Writing-Review & Editing;

## Acknowledgements

None. No funding to declare.

## Footnotes

2 ISIC-2016 (Segmentation): https://youtu.be/kB0Bf5D0WsA

3 ISIC-2017 (Segmentation): https://youtu.be/m3u58LN9lns

4 ISIC-2018 (Segmentation): https://youtu.be/r4hxv8WdQHM

5 ISIC-2016 (Classification): https://youtu.be/wwHwkQmigqU

6 ISIC-2017 (Classification): https://youtu.be/1Dn5l4g4h6Y

7 ISIC-2018 (Classification): https://youtu.be/NXVw2cyqd6k

8 https://github.com/kamruleee51/Web-App-of-Skin-Lesion-Classification

